# Is occupational physical activity associated with all-cause mortality in UK Biobank?

**DOI:** 10.1101/2020.12.18.20248428

**Authors:** Matthew Pearce, Tessa Strain, Katrien Wijndaele, Stephen J. Sharp, Alexander Mok, Soren Brage

## Abstract

**Objectives:** To investigate associations between occupational physical activity (OPA) and all-cause mortality.

**Methods:** From baseline (2006-2010), 452,884 UK Biobank participants (aged 40-69 years) were followed for a median 11.1 (IQR: 10.4-11.8) years. OPA was categorised by cross-tabulating degree of manual work and walking/standing work amongst those in paid employment (n=264,424), whereas categories of occupational status were used for those not in paid employment (n=188,460). Cox proportional hazards models were used to estimate hazard ratios (HR) and 95% confidence intervals (CI) for all-cause mortality by occupational category, and for working hours/week and non-occupational physical activity stratified by occupational category.

**Results:** During 4,965,616 person-years of follow-up, 23,310 deaths occurred. Compared to those in sedentary jobs, retirement was associated with lower mortality in women (HR=0.74, CI:0.68-0.81) and men (HR=0.85, CI:0.79-0.92), whereas unemployment was associated with higher mortality in men (HR=1.26, CI:1.10-1.45). There was no evidence of mortality differences by OPA category within the working population. Working <35 hours/week versus 35-40 hours/week was associated with lower mortality in both women (HR=0.86, CI:0.79-0.93) and men (HR=0.81, CI:0.75-0.88), with no interaction by OPA. Non-occupational physical activity was associated with lower mortality in both women (HR=0.90 per 5 kJ/day/kg, CI:0.84-0.96) and men (HR=0.88 per 5 kJ/day/kg, CI:0.84-0.92), with no interaction by OPA.

**Conclusion:** Work classified as having higher levels of OPA may not be as active as reported, or the types of physical activity performed in those jobs are not health-enhancing. Irrespective of OPA category or employment status, non-occupational physical activity appears to provide health benefits.

**SUMMARY BOX:** *What are the new findings?:*

- Retirement was associated with lower all-cause mortality, compared to having a sedentary job in both men and women but unemployment was only associated with higher mortality in men.
- There were no differences in mortality between categories with different levels of self-reported OPA
- Physical activity outside of work was associated with lower hazard of all-cause mortality and there was no interaction with occupational physical activity, indicating similar benefits across different jobs types.

*How might it impact on clinical practice in the future?:*

- Health professionals should be aware that occupations assumed to involve more physical activity may not be as active as reported, or the types of physical activity performed in those jobs may not be health-enhancing.
- Physical activity during leisure-time should continue to be recommended to adults of all paid and unpaid occupational categories.

## BACKGROUND

The benefits of physical activity for health are well established,[1,2] with guidelines from the UK Chief Medical Officers[3] and the World Health Organization[4] recommending the equivalent of 150 minutes of moderate-intensity aerobic physical activity each for maintenance of good physical and mental health. No distinctions are made between physical activity in leisure-time, transport, home, occupational domains; the total volume of activity is assumed to be beneficial regardless of the domain in which it was accrued. Contradictory to this advice is the suggestion that occupational physical activity (OPA) does not confer the same benefits, and may even be harmful to health.[5] One meta-analysis reported that male (but not female) workers with high level OPA were at 18% higher risk of mortality compared to those at low levels.[6] Reasons proposed for these findings include OPA being performed at lower intensities, for protracted periods, and in a static posture,[7] but the existing evidence also has limitations including the use of crude self-reported OPA measures, limited adjustment for non-occupational physical activity, and residual confounding for socio-economic status and lifestyle factors (e.g. smoking).[8,9] Prior studies within occupational strata have reported that active jobs were associated with lower morality.[10,11]

UK Biobank is a large prospective cohort study total and domain-specific physical activity data, as well as occupational variables. These data can be combined in such a way that both work category and hours can be used to characterise exposure to different types and volumes of OPA. A range of lifestyle, socio-economic and health-related variables are collected using a standardised protocol, and it is also possible to calibrate self-reported non-occupational physical activity to objective measures of physical activity using the accelerometer sub-cohort[12] to better control for physical activity outside of work. UK Biobank has sufficient sample size and accrued deaths to allow stratification by sex and occupational categories. This can better address issues of confounding patterned by occupational group with strata larger than many occupational cohorts. This presents opportunities for improving our understanding of OPA, particularly in the UK, where there are few contemporary analyses. In this study, we investigated whether occupational category and hours of work in different job categories were associated with all-cause mortality.

## METHODS

### Participants and study design

UK Biobank is an ongoing prospective cohort study of men and women aged 40-69 years residing within 25 miles of one of 22 assessment centres in England, Scotland, and Wales. Participants were identified from National Health Service general practitioner registries and invited to a baseline assessment between 2006 and 2010.[13] The study was approved by the North West Multicentre Research Ethics Committee and participants provided written informed consent. Data for the current analyses were updated on 26^th^ August 2020, containing information from 502,493 participants with baseline measures. Participant exclusions are outlined in Supplementary Figure 1.

**Figure 1.**
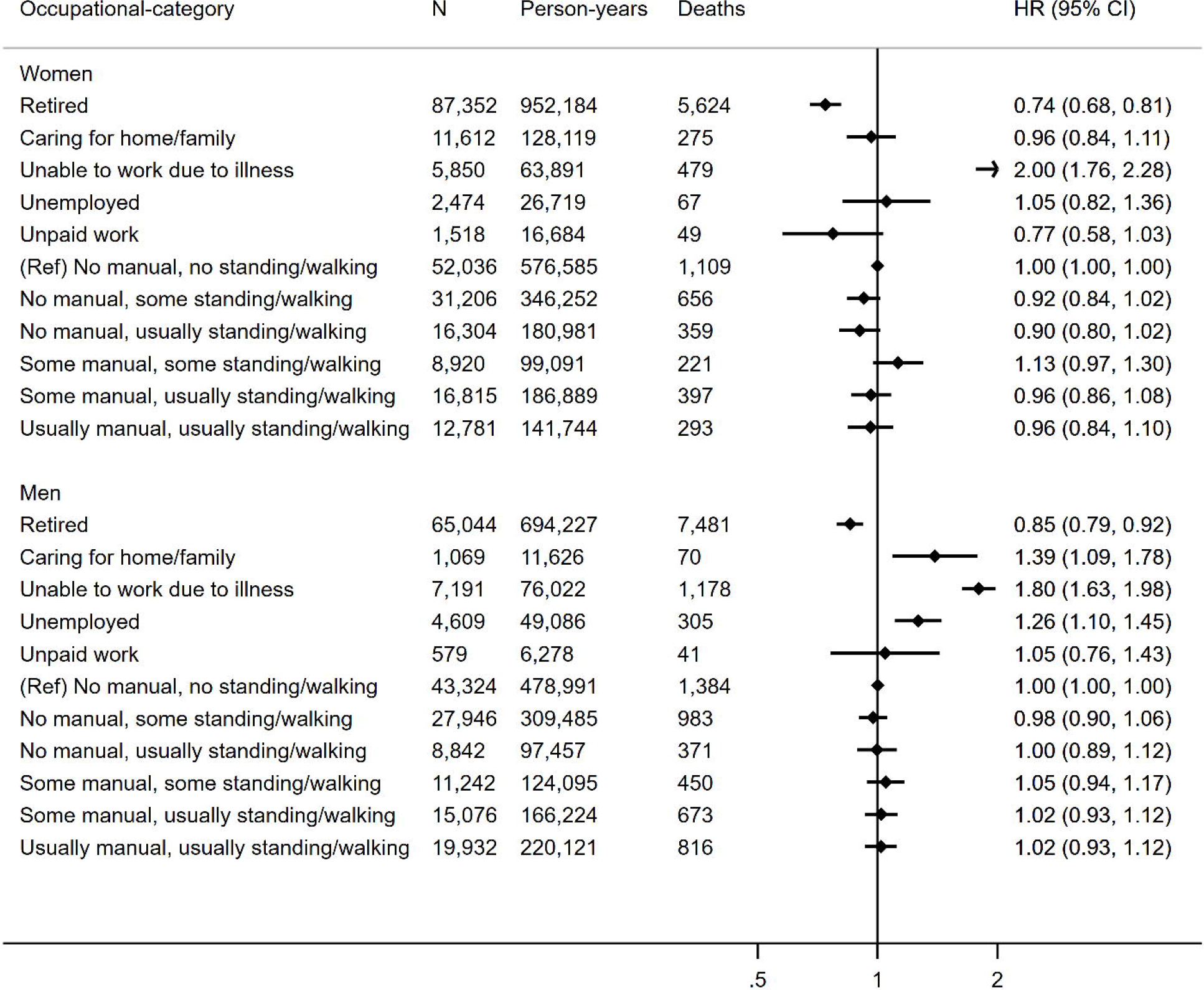
Hazard ratio (HR) and 95% confidence interval (CI) of all-cause mortality by occupational category. Reference group is “no manual, no standing/walking”. Models are adjusted for age (underlying timescale), ethnicity, Townsend deprivation index, highest educational level (stratified baseline hazard), annual household income (stratified baseline hazard), working hours per week, alcohol consumption, smoking, salt added to food, oily fish intake, fruit and vegetable intake (stratified baseline hazard), processed and red meat intake, non-occupational physical activity energy expenditure, parental history of cancer or CVD, use of blood pressure or cholesterol lowering medications, doctor-diagnosed diabetes or treatment with insulin, baseline prevalent coronary heart disease, stroke, or cancer. Data for students not shown due to small numbers.

### Exposure variables

Occupational status, standard occupational job code (SOC),[14] degree of manual work, degree of standing/walking work, and working duration in hours per week were self-reported using a touch-screen questionnaire.[15] Participants with missing data for these variables were excluded from analyses (n=9,362), as were those reporting paid employment status but zero working hours (n=186). For those in paid employment, degree of manual work and degree of standing/walking work were both reported as one of four categories: “never/rarely”, “sometimes”, “usually”, “always”. Responses of “usually” and “always” were collapsed for both manual work and standing/walking, with the two variables cross-tabulated (Supplementary Table 1) to create six mutually exclusive OPA categories: “no manual, no standing/walking”, “no manual, some standing/walking”, “no manual, usually standing/walking”, “some manual, some standing/walking”, “some manual, usually standing/walking”, “usually manual, usually standing/walking”. Total physical activity estimated by median wrist acceleration showed a predictable association across these categories within the accelerometery sub-cohort,[16] indicating face validity (Supplementary Figure 2).

**Table 1.** Baseline characteristics of women and men in UK Biobank.

|  | No paid employment |  | In paid employment |  | All |  |
| --- | --- | --- | --- | --- | --- | --- |
|  | Women | Men | Women | Men | Women | Men |
| n (%) | 109,591<br>(58) | 78,869<br>(42) | 138,062<br>(52) | 126,362<br>(48) | 247,653<br>(55) | 205,231<br>(45) |
| Working hours per week, median (IQR) | 0 (0; 0) | 0 (0; 0) | 35<br>(22;40) | 40<br>(37;45) | 15 (0;36) | 15 (0;36) |
| Age in years at baseline, mean (SD) | 61 (6) | 62 (6) | 52 (7) | 53 (7) | 56 (8) | 57 (8) |
| White ethnicity, % | 96 | 96 | 95 | 95 | 95 | 95 |
| *Highest educational level* |  |  |  |  |  |  |
| No qualification, % | 26 | 26 | 8 | 10 | 16 | 16 |
| Any other qualification, % | 50 | 45 | 53 | 51 | 52 | 49 |
| Degree or above, % | 24 | 29 | 38 | 39 | 32 | 35 |
| Townsend index (higher for more deprived), median (IQR) | -2.4<br>(-3.8;0.1) | -2.3<br>(-3.7;0.5) | -2.1<br>(-3.6;0.4) | -2.2<br>(-3.7;0.3) | -2.2<br>(-3.7;0.3) | -2.2<br>(-3.7;0.4) |
| *Household income before tax* |  |  |  |  |  |  |
| Prefer not to answer, % | 14 | 9 | 8 | 6 | 10 | 7 |
| Do not know, % | 9 | 3 | 3 | 1 | 5 | 2 |
| <£18,000, % | 31 | 35 | 11 | 7 | 20 | 18 |
| £18,000-£30,999, % | 24 | 27 | 21 | 19 | 22 | 22 |
| £31,000-£51,999, % | 13 | 17 | 28 | 30 | 22 | 25 |
| £52,000-£100,000, % | 6 | 7 | 24 | 30 | 16 | 21 |
| >£100,000, % | 2 | 1 | 6 | 8 | 4 | 6 |
| *Occupational category* |  |  |  |  |  |  |
| Retired, % | 80 | 82 |  |  | 35 | 32 |
| Caring for home/family, % | 11 | 1 |  |  | 5 | 1 |
| Unable to work due to illness, % | 5 | 9 |  |  | 2 | 4 |
| Unemployed, % | 2 | 6 |  |  | 1 | 2 |
| Unpaid work, % | 1 | 1 |  |  | 1 | 0 |
| Student, % | 1 | 0 |  |  | 0 | 0 |
| No manual, no standing/walking, % |  |  | 38 | 34 | 21 | 21 |
| No manual, Some standing/walking, % |  |  | 23 | 22 | 13 | 14 |
| No manual, Mostly standing/walking, % |  |  | 12 | 7 | 7 | 4 |
| Some manual, some standing/walking, % |  |  | 6 | 9 | 4 | 5 |
| Some manual, mostly standing/walking, % |  |  | 12 | 12 | 7 | 7 |
| Mostly manual, mostly standing/walking, % |  |  | 9 | 16 | 5 | 10 |
| *Smoking status* |  |  |  |  |  |  |
| Never, % | 58 | 42 | 61 | 54 | 60 | 50 |
| Previous, % | 34 | 46 | 30 | 34 | 32 | 39 |
| Current, % | 8 | 12 | 9 | 12 | 9 | 12 |
| *Alcohol use status* |  |  |  |  |  |  |
| Never, % | 7 | 3 | 4 | 2 | 5 | 2 |
| Previous, % | 4 | 4 | 3 | 3 | 3 | 3 |
| Current, % | 89 | 93 | 93 | 95 | 91 | 94 |
| Fruit/vegetable score, median (IQR) | 2 (1;3) | 1 (1;2) | 2 (1;3) | 1 (0;2) | 2 (1;3) | 1 (1;2) |
| Red/processed meat score, median (IQR) | 1 (1;1) | 1 (1;1) | 1 (1;1) | 1 (1;1) | 1 (1;1) | 1 (1;1) |
| Adds salt to food, % | 38 | 41 | 38 | 41 | 38 | 41 |
| Consumes oily fish, % | 28 | 31 | 35 | 38 | 32 | 35 |
| Non-work PAEE (kJ/kg/day), mean (SD) | 43 (3) | 42 (3) | 44 (3) | 43 (4) | 44 (3) | 42 (4) |
| Parental history of CVD or cancer, % | 76 | 72 | 69 | 66 | 72 | 68 |
| Blood pressure or cholesterol medication, % | 33 | 47 | 14 | 23 | 23 | 33 |
| Diagnosis of diabetes or insulin prescription, % | 5 | 10 | 2 | 5 | 4 | 7 |
| Prevalent CVD or cancer at baseline, % | 18 | 24 | 9 | 10 | 13 | 15 |
| *Body mass index* |  |  |  |  |  |  |
| <25 kg/m<sup>2</sup>, % | 36 | 25 | 43 | 26 | 40 | 25 |
| 25-30 kg/m<sup>2</sup>, % | 39 | 49 | 35 | 50 | 37 | 50 |
| >30 kg/m<sup>2</sup>, % | 25 | 26 | 22 | 25 | 23 | 25 |
| Resting heart rate in bpm, mean (SD) | 71 (11) | 69 (12) | 70 (10) | 68 (11) | 70 (11) | 68 (12) |

bpm=beats per minute; CVD=cardiovascular disease; IQR=interquartile range; PAEE=physical activity energy expenditure; SD=standard deviation.

**Figure 2.**
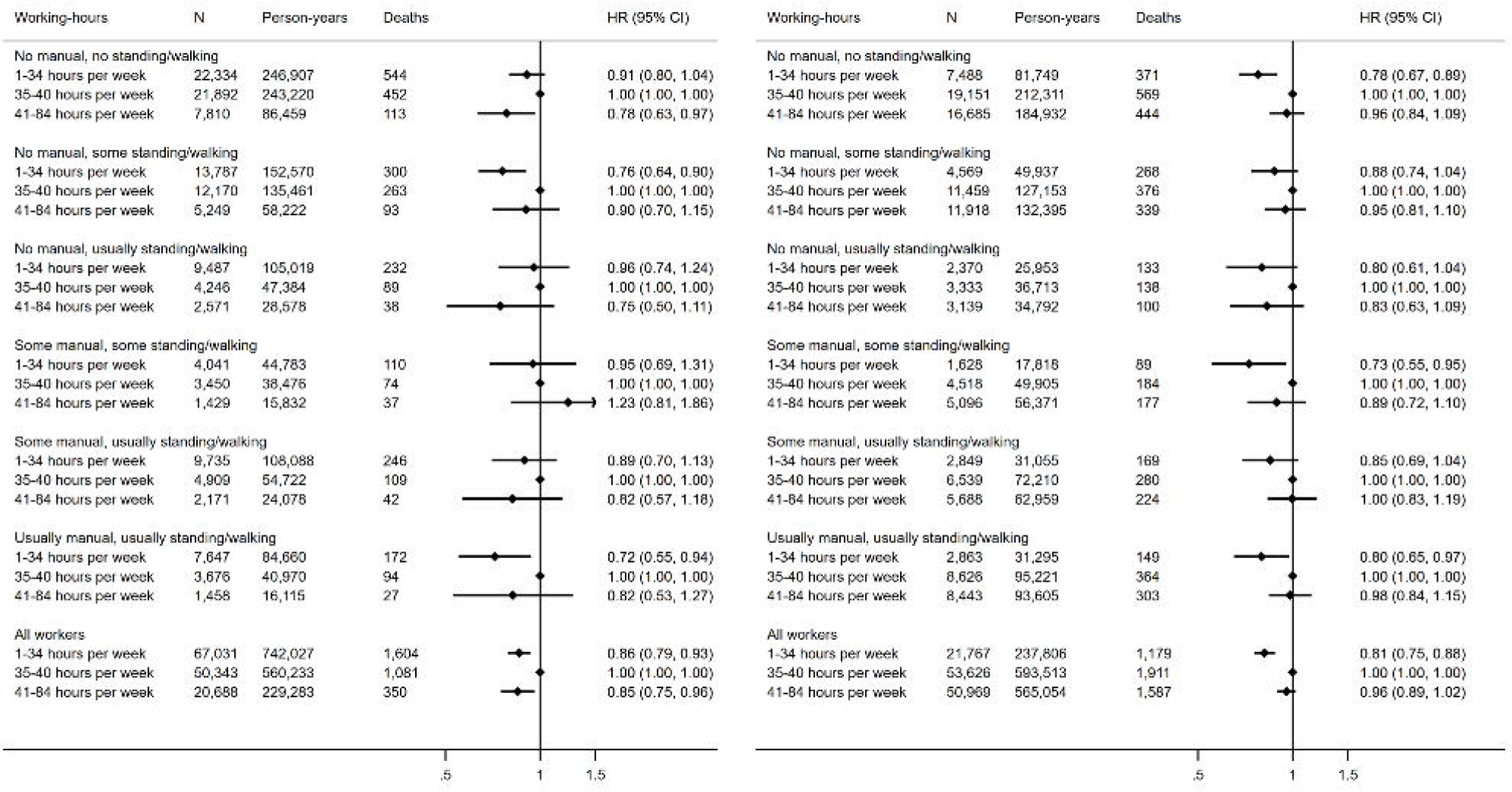
Hazard ratio (HR) and 95% confidence interval (CI) of all-cause mortality by tertile of working hours per week across occupational physical activity strata in working women (left) and men (right). Reference group is “35-40 hours per week”. Models are adjusted for age (underlying timescale), ethnicity, Townsend deprivation index, highest educational level (stratified baseline hazard), annual household income (stratified baseline hazard), alcohol consumption, smoking, salt added to food, oily fish intake, fruit and vegetable intake (stratified baseline hazard), processed and red meat intake, non-occupational physical activity energy expenditure, parental history of cancer or CVD, use of blood pressure or cholesterol lowering medications, doctor-diagnosed diabetes or treatment with insulin, baseline prevalent coronary heart disease, stroke, or cancer.

The first set of analyses used as the exposure (hereafter defined as “occupational categories”) OPA categories for those in paid employment, and occupational status of those not in paid employment (retired, unable to work due to illness/disability, caring for home/family, student, unemployed, unpaid work). The second set of analyses included only those in paid employment with tertiles of working hours as the exposure (<35, 35-40, >40 hours per week) and stratification by OPA category. In supplementary analyses we repeated both of the above but OPA category was replaced by SOC group for those in paid employment while including the same occupational status categories for those not in paid employment. Further details on the SOC group classifications are provided in Supplementary Table 2.

To investigate whether OPA moderates the association of physical activity outside of work with mortality, we examined the association between non-occupational physical activity energy expenditure (PAEE) and all-cause mortality in two sets of stratified analyses (by OPA categories and by tertiles of working hours). We previously showed how self-reported variables representing multiple behaviours could be combined to predict total PAEE in UK Biobank.[12] For the present study, the three occupational activity variables (manual work, standing/walking, sedentary work) included in this prediction model were set to zero such that the resulting estimate represented non-occupational PAEE. Further details of this prediction model are shown in Supplementary Table 3. Hazard ratios for non-occupational PAEE were presented per 5 kJ/day/kg as this approximates the lower World Health Organization guideline of 150 minutes of moderate intensity activity per week[17].

### Outcome assessment

Vital status and date of death were established by linkage to national death registries obtained from the Health and Social Care Information Centre for England and Wales and the Information Services Department for Scotland.[13] The censoring date for mortality was 31^st^ March 2020 in all three nations.

### Covariate assessment

Demographic, lifestyle, and clinical variables were assessed at baseline by the aforementioned touch-screen questionnaire, verbal interview, or physical measurement. The following baseline variables were considered as potential confounders of the relationship between OPA and all-cause mortality: age (years), sex, ethnicity (white/non-white), Townsend deprivation index (higher scores indicating higher levels of deprivation), highest educational level (degree or above/any other qualification/no qualification), annual household income (prefer not to answer, do not know, <£18,000, £18,000-£30,999, £31,000-£51,999, £52,000-£100,000, >£100,000), working hours per week (when not assessed as an exposure or used as a stratifying variable; tertiles), alcohol consumption (never/previous/current), smoking (never/previous/current), salt added to food (never/sometimes/usually), oily fish intake (never/sometimes), fruit and vegetable intake (score from 0-4 with 1 point for ≥2 servings/day for each of fresh fruit, dried fruit, cooked vegetable, raw vegetable), processed and red meat intake (average weekly frequency in days per week), parental history of cancer or CVD (yes/no), use of blood pressure or cholesterol lowering medications (yes/no), doctor-diagnosed diabetes or treatment with insulin (yes/no), baseline coronary heart disease, stroke or cancer (self-reported or ICD-10 code I20-25, I60-69, or C00-99; yes/no), body mass index (BMI) in three categories (<25, 25-30, >30 kgm^-2^), resting heart rate (beats per minute), and non-occupational PAEE (when not assessed as an exposure; kJ/day/kg).

### Statistics

For each exposure, a Cox proportional hazards model with age as the underlying timescale was used to estimate hazard ratios (HR) and 95% confidence intervals (CI) for all-cause mortality. The proportional hazards assumption for categorical variables was examined using log-log plots; the baseline hazard function was stratified by levels of those variables that did not satisfy this assumption (fruit and vegetable intake, income, education). Fractional polynomials showed that working hours per week did not meet the log-linear assumption so this variable was categorised using tertile boundaries, while meat consumption (log[x+1]) and Townsend index ([x]^2^) were transformed. Variance inflation factors and Pearson correlations indicated no strong collinearity between variables. All analyses were stratified by sex a priori, based on findings from previous studies.[6] Wald tests were used to examine the potential interactions of working hours and non-occupational PAEE with OPA category. Models included all covariates listed above, as well as separately omitting BMI and resting heart rate which may be on the causal pathway between physical activity and mortality. Individuals with missing covariate data (n=37,965) were excluded, as were those who died in the first two years of follow-up (n=2,096) to mitigate potential reverse causation. For the same reason, we conducted sensitivity analyses excluding those with prevalent coronary heart disease, stroke, or cancer at baseline (n=63,755). All analyses were performed using STATA/SE 16.1 (StataCorp, TX, USA).

### Patient and public involvement

Patients and members of the public were not formally involved in the design, analysis or interpretation of this study.

## RESULTS

In a sample of 452,884 participants, 23,310 deaths occurred during a median 11.1 (IQR: 10.4-11.8) years of follow up (4,965,616 person-years). Baseline characteristics of the participants by sex and occupational status are shown in Table 1, and further details for paid and unpaid occupational groups are provided in Supplementary Tables 4 to 7. Approximately one third of the participants were retired. Those in paid employment were younger, reported lower frequency of medication usage and had a lower prevalence of diabetes, CVD, or cancer at baseline. There were only minor differences between those in, and not in, paid employment with respect to resting heart rate, BMI, and lifestyle variables. Jobs involving no manual work or standing/walking were most common (women 38%; men 34%), whereas jobs involving the highest levels of manual work and standing/walking were less common (women 9%; men 16%). Supplementary Table 8 shows the distribution of participants across OPA categories within SOC strata. Participants in managerial, professional, and administrative SOCs tended to report less manual work and standing/walking, whereas participants in elementary, skilled trade, personal service, and operative SOCs tended to report more manual work.

Figure 1 shows hazard ratios and 95% confidence intervals of all-cause mortality for occupational categories compared with the referent of a paid job involving no manual work or standing/walking (e.g. sedentary office work). Among women and men in paid work, there were no differences in hazard of all-cause mortality. The hazard of mortality was lower in retired women, but twice as high in women unable to work due to illness. The hazard was higher in men unable to work due to illness, unemployed men, and in men caring for home or others. Additional adjustment for resting heart rate and BMI did not alter these findings (Supplementary Figure 3). When OPA category was replaced with SOC group (Supplementary Figure 4), men with “elementary” or “process, plant or machine operative” SOCs had higher hazards of all-cause mortality than those in “senior managerial positions” (the category we assumed to be most similar to sedentary desk work with large numbers in both sexes), however no such associations were observed in women. Similar associations were observed in the model adjusting for resting heart rate and BMI (Supplementary Figure 4).

**Figure 3.**
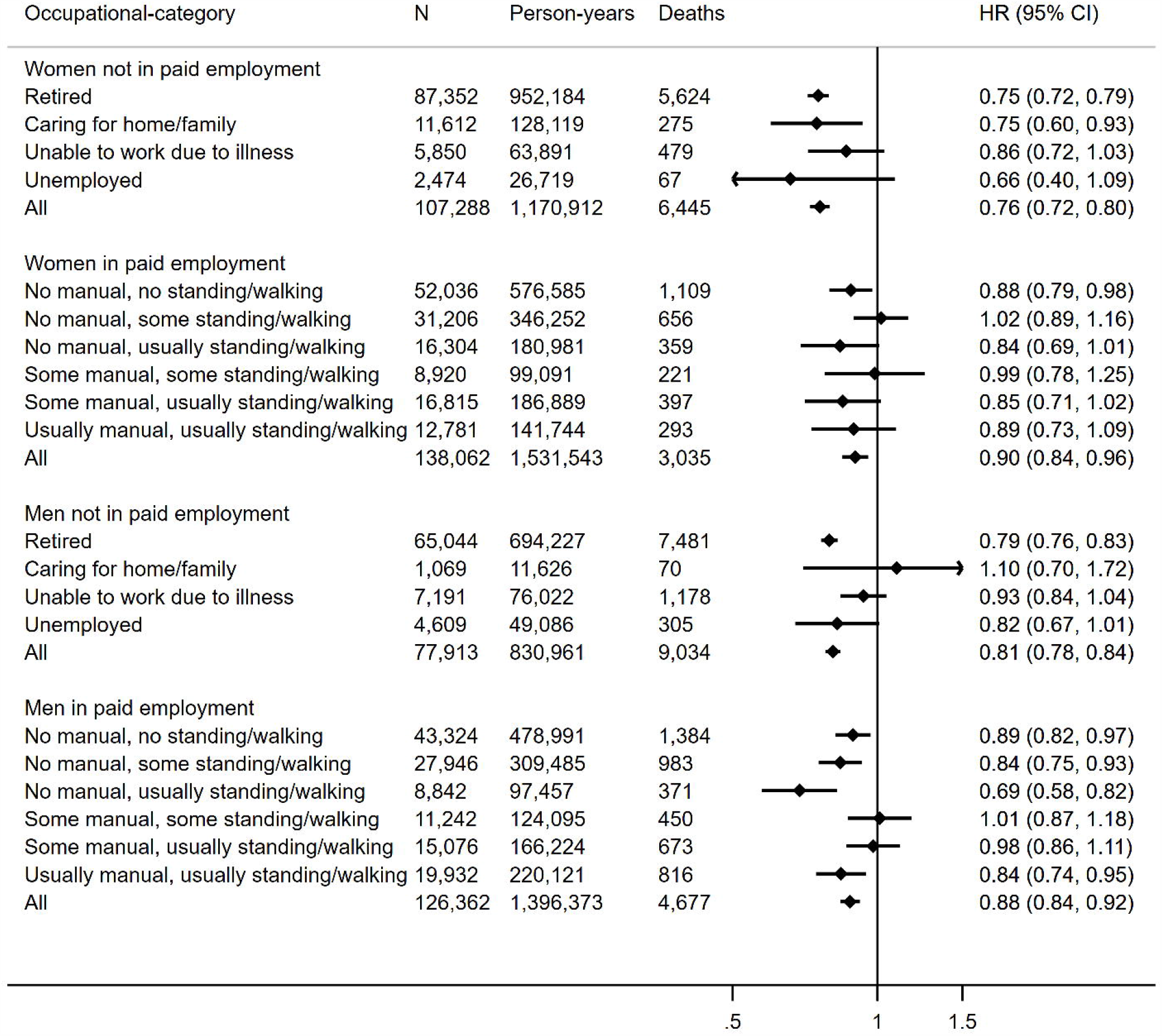
Hazard ratio (HR) and 95% confidence interval (CI) of all-cause mortality per 5 kJ/day/kg of non-occupational physical activity energy expenditure across occupational strata. Models adjusted for age (underlying timescale), ethnicity, Townsend deprivation index, highest educational level (stratified baseline hazard), annual household income (stratified baseline hazard), working hours per week, alcohol consumption, smoking, salt added to food, oily fish intake, fruit and vegetable intake (stratified baseline hazard), processed and red meat intake, parental history of cancer or CVD, use of blood pressure or cholesterol lowering medications, doctor-diagnosed diabetes or treatment with insulin, baseline prevalent coronary heart disease, stroke, or cancer. Results for students and unpaid workers not shown due to small numbers.

**Figure 4.**
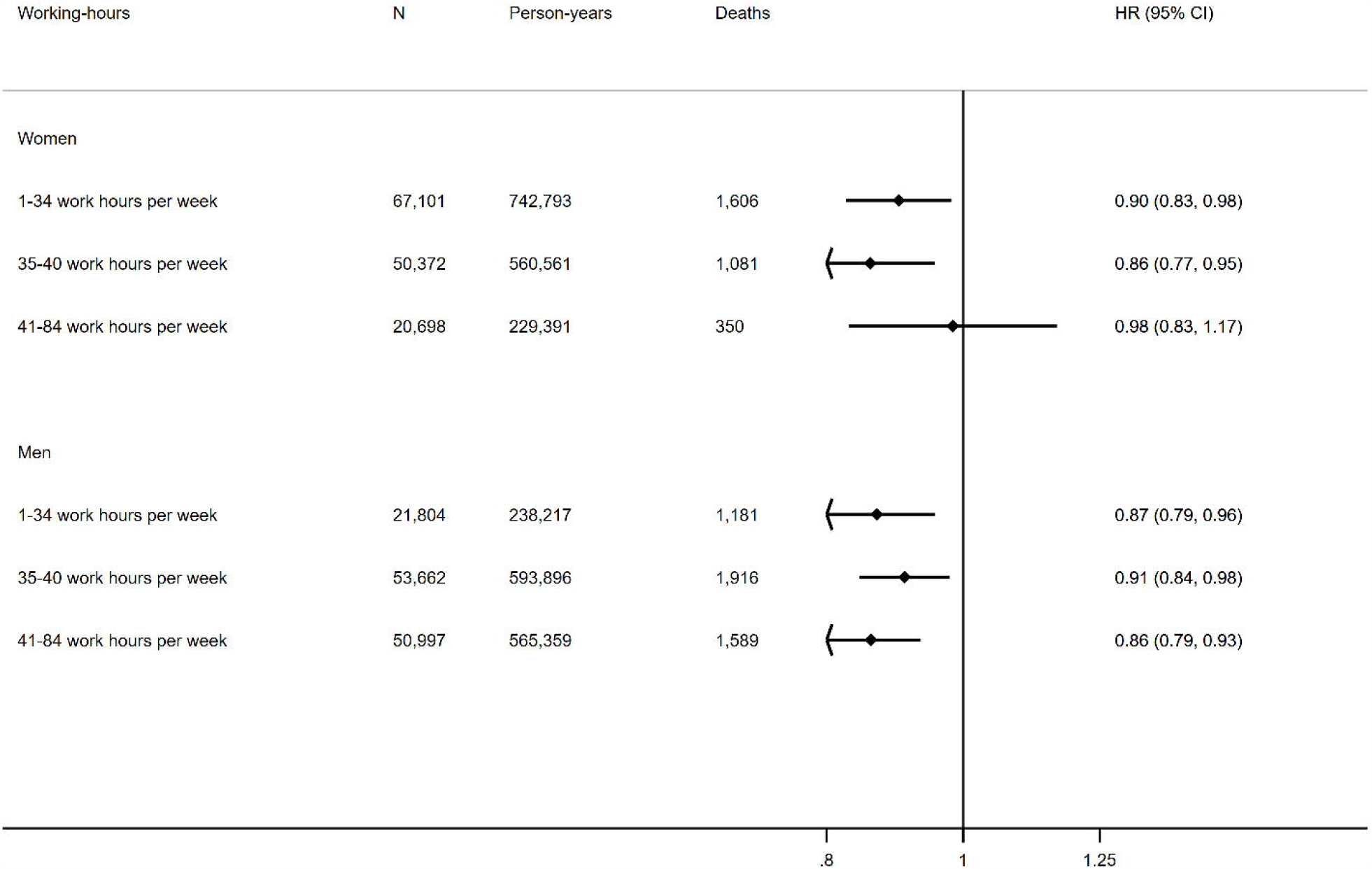
Hazard ratio (HR) and 95% confidence interval (CI) of all-cause mortality per 5 kJ/day/kg of non-occupational physical activity energy expenditure across tertiles of working hours per week. Models adjusted for age (underlying timescale), ethnicity, Townsend deprivation index, highest educational level (stratified baseline hazard), annual household income (stratified baseline hazard), alcohol consumption, smoking, salt added to food, oily fish intake, fruit and vegetable intake (stratified baseline hazard), processed and red meat intake, parental history of cancer or CVD, use of blood pressure or cholesterol lowering medications, doctor-diagnosed diabetes or treatment with insulin, baseline prevalent coronary heart disease, stroke, or cancer, occupational physical activity category.

Figure 2 shows hazards of all-cause mortality for tertiles of working hours per week within different OPA strata. Women working 35-40 hours per week had higher hazard than those working 1-34 hours per week, but women working the longest hours had lower hazard than those in the middle tertile. Among men, working 35-40 or >40 hours per week was associated with similarly high hazards of mortality, compared with those working 1-34 hours per week. There was no evidence of interaction between working hours and OPA category (p=0.49 and p=0.90 for women and men, respectively). Additional adjustment for resting heart rate and BMI did not materially alter these findings (Supplementary Figure 5). There was no evidence of interaction between working hours and SOC category (p=0.32 and p=0.68 for women and men, respectively; Supplementary Figure 6).

Figure 3 shows associations between non-occupational PAEE and all-cause mortality across occupational strata. For those in paid employment, non-occupational PAEE was associated with lower hazard of mortality in both sexes with no evidence of interaction by occupational group (p=0.19 and p=0.11 for women and men, respectively). For those not in paid employment, non-occupational PAEE was also associated with lower hazard of mortality in both sexes with evidence of interaction by occupational group in men (p=0.02) but not women (p=0.40). Following additional adjustment for resting heart rate and BMI, hazard ratios were attenuated across all strata (Supplementary Figure 7).

The inverse association between non-occupational PAEE and mortality was reasonably consistent across tertiles of working hours with no evidence of interaction observed in either women (p=0.69) or men (p=0.61) (Figure 4). Following additional adjustment for resting heart rate and BMI, hazard ratios were attenuated across all strata (Supplementary Figure 8). We observed similar results when repeating all of the above analyses with the exclusion of participants with baseline prevalent coronary heart disease, stroke or cancer (data not shown).

## DISCUSSION

In this study of 452,884 women and men including 264,424 paid workers in occupations with varying degrees of manual work and standing/walking, we found little evidence that all-cause mortality varied by category of OPA. Working full-time rather than part-time hours was associated with higher hazard of mortality but there was no pattern indicating that hours in some OPA categories were more harmful than others. Retirement was associated with lower mortality in both men and women but not working due to illness at baseline was predictably not beneficial for survival. Non-occupational physical activity was beneficial across occupational categories, supporting universal physical activity guidelines.[3]

We found no evidence of an association between OPA and all-cause mortality after controlling for non-occupational physical activity, working hours, and a range of demographic, clinical, and lifestyle variables. This is somewhat in contrast to a meta-analysis of 193,696 people reporting that men with high level of OPA were at higher risk of mortality than those at the low level (HR=1.18, 95% CI: 1.05-1.34, I^2^=76%), and the corresponding result for women which showed some evidence of an inverse association (HR=0.90, 95% CI: 0.80-1.01, I^2^=0%).[6] Our main findings from a single UK cohort are however in agreement with studies from Europe[18–22] and the USA[23] indicating no association. However, in our SOC analysis, we did observe higher all-cause mortality for “elementary occupations” and “process, plant and machine operatives” in men. Discrepancies between our findings and previous work may partly be explained by variation in working patterns and conditions between populations and eras. For example, the strongest effect size (HR 3.40, 95% CI: 1.94-5.96) in the above meta-analysis is from a Taiwanese study with baseline in 1990,[24] likely not generalisable to the UK between 2006 and 2010. Alternatively, our findings could suggest that in this UK population, the combination of two self-reported OPA variables is insufficient to characterise the intensity level of work throughout the day or week, making groups more difficult to distinguish and biasing effect estimates towards the null. The SOCs for which we observed higher harmful associations with all-cause mortality (including assembly line and construction workers, cleaners, and drivers) are perhaps more consistent in terms of activity intensity and thus better characterised, but the potential risks of the actual physical activity performed as part of these occupations should be investigated further using objective measures of physical activity labelled by domain. Accelerometers have been combined with work diaries to show that for mostly (71%) “blue-collar” workers in Denmark, reallocating time to MVPA at work was positively associated with long term sickness absence, whereas an inverse association was observed for reallocating time to MVPA in leisure-time.[25]

In contrast to objective monitoring, self-reported categorical data do not detail the pattern of work bouts intensity across each day. Although a strength of this work was calibration of our estimate of non-occupational PAEE to objective measures, we have previously shown that the combined inference of activity volume from these self-report data is weak relative to objective measures, with a large proportion of unexplained variance typical of self-report data.[26] This unexplained variance would include unmeasured OPA which could vary by occupation. Without methods to more accurately estimate the dose of OPA and control for non-occupational activity, it is at present difficult to rule out the possibility of health risks or benefits associated with OPA, let alone make domain-specific health recommendations such as those relating to total physical activity, for which robust measures are available.[27]

Overall, we found that non-occupational physical activity was inversely associated with all-cause mortality in paid workers, with stronger associations for those not in paid work. We observed no interaction between OPA category and non-occupational PAEE in paid workers, reflecting previous reports that leisure-time physical activity was beneficial independent of occupational physical activity level.[28,29] Taken together, our results for OPA and non-occupational physical activity suggest that all adults should all aim to be active during their leisure-time irrespective of their occupational status, with the potential additional benefit of substituting out harmful sedentary behaviours.[30] Our results also indicate that OPA may not confer health benefits in this relatively older UK population so the message to be active in leisure-time may be even more important. Moreover, increasing activity at work may be difficult for some workers.

Strengths of this work include a large single cohort study allowing robust estimation of associations, adjustment for a wide range of socio-economic and behavioural covariates as well as sufficient size to conduct stratified analyses larger than many occupational cohort studies. There are also important limitations of this work. As in any observational study, the above adjustments cannot fully eliminate confounding. Job satisfaction, exposure to hazardous materials or working conditions, and shift pattern data are available in UK Biobank but only in a small subsample. Data for phenomena which may potentially confound associations, such as work stress[31,32] and access to sick leave[33] are not recorded. Characteristics like these may be patterned by occupational group, and these strata could be used to investigate specific working cohorts, such as in previous studies.[10,11]

We used baseline data to assign occupational status and were unable to account for any changes during the follow-up period. In a sample aged 40-69 at baseline, retirement or changes in work due to illness during follow-up are of particular concern. We were also unable to account for potentially complex work histories leading up to baseline, account for changes to nature of work over time,[34] or generalise our findings to younger workers. There is also evidence of a healthy volunteer selection bias in UK Biobank such that it is not representative of the general population,[35] particularly in relation to smoking and education[36] which are notable confounders for this study.

In summary, analysis of this population of UK adults aged 40-69 years old showed limited evidence of an association between OPA and all-cause mortality, although potential measurement error and residual confounding mean that we are unable to rule out the possibility of either health benefits or risks. Until stronger evidence is available from a combination of domain labels and objective assessment of the temporal pattern of activity, individuals should continue to maximise their physical activity volume during leisure-time irrespective of their occupation.

## Contributors

MP and SB conceptualised the study. MP undertook the analyses with input from TS, SJS, KW, AM, and SB. MP drafted the manuscript with critical revisions from all authors. All authors approved the final version.

The lead author (MP) affirms that this manuscript is an honest, accurate, and transparent account of the study being reported; that no important aspects of the study have been omitted; and that any discrepancies from the study as planned have been explained.

## Supporting information

STROBE checklist

## Data Availability

The UK Biobank data that support the findings of this study are available to all bona fide researchers for health related research that is in the public interest, https://www.ukbiobank.ac.uk/register-apply/. This work was conducted under UK Biobank application number 20684.

## Acknowledgements

We are indebted to the principal investigators of UK Biobank and the volunteers who took part.

## Funding

MP, TS, KW, AM, and SB acknowledge funding from the Medical Research Council (grants MC_UU_00006/4, MC_UU_12015/3, and MC_UU_12015/1) and NIHR Cambridge Biomedical Research Centre (IS_BRC-1215-20014). SJS acknowledges funding from MRC grant MC_UU_12015/1.

### Ethical approval

UK Biobank was approved by the North West Multicentre Research Ethics Committee and all participants provided written informed consent.

