## Supplementary data for "Is occupational physical activity associated with all-cause mortality in UK Biobank?"

**Table S1** Creation of joint standing/walking work and manual work variable.

|  |  | Heavy manual or physical | | |
| --- | --- | --- | --- | --- |
|  |  | Never/rarely | Sometimes | Mostly/always |
| Standing/walking | Never/rarely | Group 1 (n=95,040) | Group 1 (n=4,247) | Group 1  (n=628) |
|  | Sometimes | Group 2  (n=62,382) | Group 4  (n=21,812) | Group 6  (n=2,746) |
|  | Mostly/always | Group 3  (n=26,897) | Group 5  (n=34,597) | Group 6  (n=33,204) |

All participants reporting never/rarely walking or standing at work were included in category 1 on the basis that it would be unusual to be involved in heavy manual work without standing. There were few participants reporting sometimes walking or standing but mostly/always being involved in heavy manual work, so these were included in category 6 on the basis that they would be similar to that group given their higher frequency of heavy manual work. Heavy manual or physical work from data field 816 (<http://biobank.ctsu.ox.ac.uk/crystal/field.cgi?id=816>), standing/walking work from data field 806 (<http://biobank.ctsu.ox.ac.uk/crystal/field.cgi?id=806>).

**Table S2** Standard occupational codes (SOC) of paid workers in UK Biobank.

| First level SOC | Types of occupation |
| --- | --- |
| Managers and senior officials | Corporate managers and senior officials |
|  | Production managers |
|  | Functional managers |
|  | Quality and customer care managers |
|  | Financial institution and office managers |
|  | Managers in distribution, storage and retailing |
|  | Protective service officers |
|  | Health and social services managers |
|  | Managers in farming, horticulture, forestry and fishing |
|  | Managers and proprietors in hospitality and leisure services |
|  | Managers and proprietors in other service industries |
| Professional occupations | Science professionals |
|  | Engineering professionals |
|  | Information and communication technology professionals |
|  | Health professionals |
|  | Teaching professionals |
|  | Research professionals |
|  | Legal professionals |
|  | Business and statistical professionals |
|  | Architects, town planners, surveyors |
|  | Public service professionals |
|  | Librarians and related professionals |
| Associate professional and technical occupations | Science and engineering technicians |
|  | Draughtspersons and building inspectors |
|  | IT service delivery occupations |
|  | Health associate professionals |
|  | Therapists |
|  | Social welfare associate professionals |
|  | Protective service occupations |
|  | Artistic and literary occupations |
|  | Design associate professionals |
|  | Media associate professionals |
|  | Sports and fitness occupations |
|  | Transport associate professionals |
|  | Legal associate professionals |
|  | Business and finance associate professionals |
|  | Sales and related associate professionals |
|  | Conservation associate professionals |
|  | Public service and other associate professionals |
| Administrative and secretarial occupations | Administrative occupations: government and related organisations |
|  | Administrative occupations: finance |
|  | Administrative occupations: records |
|  | Administrative occupations: communications |
|  | Administrative occupations: general |
|  | Secretarial and related occupations |
| Skilled trades occupations | Agricultural trades |
|  | Metal forming, welding and related trades |
|  | Metal machining, fitting and instrument making trades |
|  | Vehicle trades |
|  | Electrical trades |
|  | Construction trades |
|  | Building trades |
|  | Textiles and garments trades |
|  | Printing trades |
|  | Food preparation trades |
| Personal service occupations | Healthcare and related personal services |
|  | Childcare and related personal services |
|  | Animal care services |
|  | Leisure and travel service occupations |
|  | Hairdressers and related occupations |
|  | Housekeeping occupations |
| Sales and customer service occupations | Sales assistants and retail cashiers |
|  | Sales related occupations |
|  | Customer service occupations |
| Process, plant and machine operatives | Process operatives |
|  | Plant and machine operatives |
|  | Assemblers and routine operatives |
|  | Construction operatives |
|  | Transport drivers and operatives |
|  | Mobile machine drivers and operatives |
| Elementary occupations | Elementary agricultural occupations |
|  | Elementary construction occupations |
|  | Elementary process plant occupations |
|  | Elementary goods storage occupations |
|  | Elementary administration occupations |
|  | Elementary personal services occupations |
|  | Elementary cleaning occupations |
|  | Elementary security occupations |
|  | Elementary sales occupations |

Standard occupational codes from data field 20277 (<http://biobank.ctsu.ox.ac.uk/crystal/field.cgi?id=20277>)

**Table S3** Mutually adjusted sex-specific coefficients (standard errors) for prediction of average daily wrist acceleration (ln milli-g) from 13 self-reported behaviours.

| Variable | Women (n=52,507) | Men (n=40,918) |
| --- | --- | --- |
| Walking for pleasure (ln min/day) | .0227 (.0009) | .0177 (.0010) |
| Strenuous sports (ln min/day) | .0377 (.0014) | .0416 (.0014) |
| Other exercises (ln min/day) | .0242 (.0008) | .0280 (.0009) |
| Light DIY activities (min/day) | .0018 (.0009) | .0043 (.0010) |
| Heavy DIY activities (ln min/day) | .0156 (.0011) | .0163 (.0011) |
| Heavy physical work (ln min/day) | .0120 (.0009) | .0189 (.0010) |
| Standing/walking at work (ln min/day) | .0099 (.0006) | .0095 (.0007) |
| Sedentary time at work (ln min/day) | .0039 (.0005) | .0048 (.0006) |
| Getting about method |  |  |
| Car or public transportation | Reference | Reference |
| Mixed use | .0080 (.0025) | .0112 (.0030) |
| Walking or cycling | .0360 (.0044) | .0502 (.0053) |
| Commuting method |  |  |
| Car or public transportation | Reference | Reference |
| Mixed use | .0366 (.0039) | .0391 (.0047) |
| Walking or cycling | .0522 (.0054) | .0539 (.0071) |
| TV viewing (hours/day) | -.0827 (.0026) | -.0817 (.0033) |
| Computer use (ln hours/day) | -.0609 (.0027) | -.0639 (.0030) |
| Sleep |  |  |
| ≤5.0 (hours/day) | -.0254 (.0064) | -.0080 (.0082) |
| 6.0 (hours/day) | -.0132 (.0032) | .0045 (.0038) |
| 7.0 (hours/day) | Reference | Reference |
| 8.0 (hours/day) | -.0206 (.0027) | -.0305 (.0034) |
| ≥9.0 (hours/day) | -.0735 (.0049) | -.0873 (.0064) |
| Constant | 3.378 (.0082) | 3.327 (.0102) |

To estimate non-occupational physical activity energy expenditure (PAEE), we used a method similar to one previously described [<https://doi.org/10.1186/s12966-020-00937-4>]. Briefly, the previous method generated sex-specific regression models developed in the UK Biobank accelerometery sub-cohort to predict mean wrist acceleration from 14 self-reported behaviours. These self-reported behaviours and regression coefficients were then used to predict wrist acceleration in the main UK Biobank cohort. Predicted wrist acceleration was then converted to total daily PAEE in kJ/day/kg using data from a similarly aged UK cohort [<https://dx.doi.org/10.1371%2Fjournal.pone.0167472>] and a previously reported scaling equation for dominant wrist acceleration [<https://doi.org/10.1038/s41366-019-0352-x>].

For the present study, there were two adjustments to the above method for the slightly different purpose of predicting non-occupational PAEE. Firstly, we excluded moderate-to-vigorous physical activity (MVPA) from the prediction model because the questions used for this variable included both leisure and work activity, and so any subsequent prediction of non-occupational PAEE might include work related activity. This meant that 13 rather than 14 self-reported behaviours were used to generate the sex-specific regression models for predicting wrist acceleration, as shown above. The newly derived regression models and the original models including MVPA explained 14% and 17% variance in wrist acceleration for women and men respectively (i.e. there was no noticeable change in explanatory power when excluding MVPA).

Secondly, although still included in the prediction model, variables representing occupational physical activity (heavy manual work, standing/walking work, sedentary work) were set to zero at the prediction stage so that the resulting estimate represented non-occupational PAEE. Natural log transformations of self-reported behaviours used ln(x+1).

**Table S4** Baseline characteristics of women in paid employment in UK Biobank.

|  | No manual, no standing/walking | No manual, some standing/walking | No manual, usually standing/walking | Some manual, some standing/walking | Some manual, usually standing/walking | Usually manual, usually standing/walking |
| --- | --- | --- | --- | --- | --- | --- |
| n (%) | 52,036 (38) | 31,206 (27) | 16,304 (12) | 8,920 (6) | 16,815 (12) | 12,781 (9) |
| Working hours per week, median (IQR) | 35 (24 to 40) | 35 (24 to 40) | 30 (18 to 40) | 35 (24 to 40) | 30 (20 to 38) | 30 (20 to 38) |
| Age in years at baseline, mean (SD) | 52 (7) | 53 (7) | 53 (7) | 52 (7) | 53 (7) | 52 (7) |
| White ethnicity, % | 96 | 95 | 94 | 94 | 93 | 92 |
| *Highest educational level* |  |  |  |  |  |  |
| No qualification, % | 5 | 6 | 10 | 8 | 14 | 20 |
| Any other qualification, % | 53 | 50 | 44 | 57 | 60 | 63 |
| Degree or above, % | 42 | 44 | 46 | 35 | 25 | 17 |
| Townsend index (higher for more deprived), median (IQR) | -2.2 (-3.7 to 0.2) | -2.2 (-3.7 to 0.2) | -2.3 (-3.7 to 0.1) | -2.0 (-3.5 to 0.7) | -1.8 (-3.4 to 1.0) | -1.4 (-3.2 to 1.7) |
| *Household income before tax* |  |  |  |  |  |  |
| Prefer not to answer, % | 7 | 8 | 8 | 8 | 9 | 9 |
| Do not know, % | 1 | 2 | 3 | 3 | 4 | 5 |
| <£18,000, % | 7 | 8 | 12 | 13 | 18 | 24 |
| £18,000-£30,999, % | 19 | 20 | 20 | 24 | 25 | 27 |
| £31,000-£51,999, % | 29 | 30 | 29 | 29 | 27 | 23 |
| £52,000-£100,000, % | 29 | 27 | 24 | 20 | 15 | 11 |
| >£100,000, % | 8 | 6 | 4 | 3 | 2 | 2 |
| *Smoking status* |  |  |  |  |  |  |
| Never, % | 61 | 61 | 65 | 61 | 60 | 57 |
| Previous, % | 31 | 30 | 27 | 30 | 28 | 29 |
| Current, % | 8 | 8 | 8 | 10 | 12 | 14 |
| *Alcohol use status* |  |  |  |  |  |  |
| Never, % | 3 | 4 | 5 | 4 | 6 | 7 |
| Previous, % | 2 | 2 | 3 | 3 | 3 | 4 |
| Current, % | 95 | 94 | 92 | 93 | 91 | 90 |
| Fruit/vegetable score, median (IQR) | 2 (1 to 2) | 2 (1 to 3) | 2 (1 to 3) | 2 (1 to 3) | 2 (1 to 3) | 2 (1 to 3) |
| Red/processed meat score, median (IQR) | 1 (1 to 1) | 1 (1 to 1) | 1 (1 to 1) | 1 (1 to 1) | 1 (1 to 1) | 1 (1 to 1) |
| Adds salt to food, % | 37 | 38 | 36 | 40 | 40 | 40 |
| Consumes oily fish, % | 37 | 35 | 35 | 33 | 35 | 33 |
| Non-work PAEE (kJ/kg/day), mean (SD) | 44 (3) | 44 (3) | 44 (3) | 44 (3) | 44 (3) | 44 (3) |
| Parental history of CVD or cancer, % | 68 | 70 | 69 | 69 | 69 | 68 |
| Blood pressure or cholesterol medication, % | 13 | 15 | 15 | 14 | 16 | 15 |
| Diagnosis of diabetes or insulin prescription, % | 2 | 2 | 2 | 3 | 3 | 3 |
| Prevalent CVD or cancer at baseline, % | 9 | 9 | 9 | 9 | 10 | 9 |
| *Body mass index* |  |  |  |  |  |  |
| <25 kg/m2, % | 45 | 43 | 45 | 40 | 40 | 40 |
| 25-30 kg/m2, % | 34 | 36 | 36 | 35 | 37 | 36 |
| >30 kg/m2, % | 21 | 22 | 19 | 24 | 23 | 24 |
| Resting heart rate in bpm, mean (SD) | 69 (10) | 70 (10) | 70 (10) | 70 (10) | 70 (10) | 70 (10) |

bpm=beats per minute; CVD=cardiovascular disease; IQR=interquartile range; PAEE=physical activity energy expenditure; SD=standard deviation.

**Table S5** Baseline characteristics of men in paid employment in UK Biobank.

|  | No manual, no standing/walking | No manual, some standing/walking | No manual, usually standing/walking | Some manual, some standing/walking | Some manual, usually standing/walking | Usually manual, usually standing/walking |
| --- | --- | --- | --- | --- | --- | --- |
| n (%) | 43,324 (34) | 27,946 (22) | 8,842 (7) | 11,242 (9) | 15,076 (12) | 19,932 (16) |
| Working hours per week, median (IQR) | 40 (36 to 45) | 40 (37 to 45) | 40 (30 to 45) | 40 (37 to 48) | 40 (36 to 45) | 40 (37 to 48) |
| Age in years at baseline, mean (SD) | 53 (7) | 54 (7) | 54 (7) | 53 (7) | 54 (7) | 53 (7) |
| White ethnicity, % | 96 | 95 | 92 | 94 | 92 | 95 |
| *Highest educational level* |  |  |  |  |  |  |
| No qualification, % | 4 | 4 | 10 | 14 | 18 | 26 |
| Any other qualification, % | 42 | 45 | 44 | 61 | 62 | 64 |
| Degree or above, % | 54 | 51 | 46 | 25 | 20 | 10 |
| Townsend index (higher for more deprived), median (IQR) | -2.5 (-3.9 to -0.1) | -2.6 (-3.9 to -0.4) | -2.1 (-3.7 to 0.4) | -2.1 (-3.6 to 0.4) | -1.8 (-3.4 to 1.0) | -1.5 (-3.2 to 1.3) |
| *Household income before tax* |  |  |  |  |  |  |
| Prefer not to answer, % | 4 | 5 | 5 | 7 | 7 | 8 |
| Do not know, % | 1 | 1 | 1 | 1 | 2 | 3 |
| <£18,000, % | 3 | 3 | 8 | 8 | 12 | 15 |
| £18,000-£30,999, % | 12 | 12 | 20 | 22 | 29 | 34 |
| £31,000-£51,999, % | 27 | 30 | 32 | 35 | 32 | 30 |
| £52,000-£100,000, % | 39 | 38 | 28 | 24 | 16 | 10 |
| >£100,000, % | 14 | 12 | 6 | 3 | 2 | 1 |
| *Smoking status* |  |  |  |  |  |  |
| Never, % | 58 | 57 | 55 | 51 | 49 | 49 |
| Previous, % | 33 | 34 | 33 | 36 | 36 | 34 |
| Current, % | 10 | 9 | 11 | 13 | 15 | 17 |
| *Alcohol use status* |  |  |  |  |  |  |
| Never, % | 2 | 2 | 3 | 2 | 3 | 2 |
| Previous, % | 2 | 2 | 3 | 3 | 3 | 3 |
| Current, % | 96 | 96 | 93 | 95 | 94 | 94 |
| Fruit/vegetable score, median (IQR) | 1 (1 to 2) | 1 (1 to 2) | 1 (1 to 2) | 1 (0 to 2) | 1 (0 to 2) | 1 (0 to 2) |
| Red/processed meat score, median (IQR) | 1 (1 to 1) | 1 (1 to 1) | 1 (1 to 1) | 1 (1 to 1) | 1 (1 to 1) | 1 (1 to 1) |
| Adds salt to food, % | 38 | 40 | 40 | 44 | 44 | 45 |
| Consumes oily fish, % | 39 | 37 | 38 | 37 | 36 | 36 |
| Non-work PAEE (kJ/kg/day), mean (SD) | 43 (4) | 43 (4) | 43 (4) | 43 (4) | 43 (4) | 43 (4) |
| Parental history of CVD or cancer, % | 66 | 68 | 67 | 66 | 66 | 65 |
| Blood pressure or cholesterol medication, % | 22 | 25 | 26 | 24 | 25 | 22 |
| Diagnosis of diabetes or insulin prescription, % | 4 | 5 | 5 | 5 | 5 | 4 |
| Prevalent CVD or cancer at baseline, % | 9 | 10 | 11 | 10 | 11 | 9 |
| *Body mass index* |  |  |  |  |  |  |
| <25 kg/m2, % | 28 | 25 | 25 | 21 | 24 | 24 |
| 25-30 kg/m2, % | 49 | 51 | 51 | 50 | 50 | 50 |
| >30 kg/m2, % | 23 | 24 | 24 | 29 | 27 | 26 |
| Resting heart rate in bpm, mean (SD) | 68 (11) | 68 (11) | 68 (11) | 68 (11) | 68 (11) | 68 (11) |

bpm=beats per minute; CVD=cardiovascular disease; IQR=interquartile range; PAEE=physical activity energy expenditure; SD=standard deviation

**Table S6** Baseline characteristics of women not in paid employment in UK Biobank.

|  | Retired | Caring for home/family | Unable to work due to illness | Unemployed | Unpaid work | Student |
| --- | --- | --- | --- | --- | --- | --- |
| n (%) | 87,352 (80) | 11,612 (11) | 5,850 (5) | 2,474 (2) | 1,518 (1) | 785 (1) |
| Working hours per week, median (IQR) | 0 | 0 | 0 | 0 | 0 | 0 |
| Age in years at baseline, mean (SD) | 64 (4) | 52 (7) | 53 (6) | 51 (6) | 57 (7) | 48 (6) |
| White ethnicity, % | 98 | 92 | 92 | 81 | 93 | 79 |
| *Highest educational level* |  |  |  |  |  |  |
| No qualification, % | 28 | 16 | 32 | 19 | 9 | 5 |
| Any other qualification, % | 49 | 54 | 52 | 53 | 46 | 44 |
| Degree or above, % | 23 | 30 | 16 | 27 | 45 | 51 |
| Townsend index (higher for more deprived), median (IQR) | -2.5 (-3.8 to -0.3) | -2.38 (-3.8 to 0.2) | 0.7 (-2.3 to 3.7) | 0.3 (-2.4 to 3.5) | -2.1 (-3.7 to 0.6) | -0.5 (-2.8 to 2.9) |
| *Household income before tax* |  |  |  |  |  |  |
| Prefer not to answer, % | 14 | 13 | 8 | 10 | 17 | 10 |
| Do not know, % | 8 | 9 | 11 | 10 | 9 | 7 |
| <£18,000, % | 32 | 17 | 50 | 47 | 20 | 36 |
| £18,000-£30,999, % | 26 | 15 | 17 | 13 | 16 | 15 |
| £31,000-£51,999, % | 13 | 17 | 9 | 11 | 15 | 16 |
| £52,000-£100,000, % | 5 | 17 | 3 | 6 | 13 | 13 |
| >£100,000, % | 1 | 11 | 1 | 2 | 9 | 3 |
| *Smoking status* |  |  |  |  |  |  |
| Never, % | 59 | 64 | 47 | 55 | 62 | 60 |
| Previous, % | 35 | 28 | 29 | 26 | 30 | 30 |
| Current, % | 6 | 8 | 24 | 19 | 8 | 10 |
| *Alcohol use status* |  |  |  |  |  |  |
| Never, % | 6 | 8 | 11 | 11 | 7 | 7 |
| Previous, % | 4 | 4 | 13 | 5 | 4 | 5 |
| Current, % | 90 | 88 | 76 | 84 | 89 | 88 |
| Fruit/vegetable score, median (IQR) | 2 (1 to 3) | 2 (1 to 3) | 1 (1 to 2) | 1 (1 to 2) | 2 (1 to 3) | 2 (1 to 3) |
| Red/processed meat score, median (IQR) | 1 (1 to 1) | 1 (1 to 1) | 1 (1 to 1) | 1 (1 to 1) | 1 (1 to 1) | 1 (1 to 1) |
| Adds salt to food, % | 38 | 38 | 39 | 39 | 38 | 39 |
| Consumes oily fish, % | 27 | 33 | 32 | 33 | 29 | 35 |
| Non-work PAEE (kJ/kg/day), mean (SD) | 43 (3) | 44 (3) | 41 (3) | 43 (3) | 44 (3) | 43 (4) |
| Parental history of CVD or cancer, % | 77 | 68 | 75 | 65 | 74 | 61 |
| Blood pressure or cholesterol medication, % | 36 | 15 | 36 | 18 | 20 | 9 |
| Diagnosis of diabetes or insulin prescription, % | 5 | 4 | 10 | 5 | 3 | 2 |
| Prevalent CVD or cancer at baseline, % | 19 | 10 | 25 | 11 | 13 | 7 |
| *Body mass index* |  |  |  |  |  |  |
| <25 kg/m2, % | 36 | 47 | 26 | 34 | 49 | 45 |
| 25-30 kg/m2, % | 41 | 32 | 31 | 35 | 32 | 35 |
| >30 kg/m2, % | 24 | 20 | 44 | 31 | 19 | 21 |
| Resting heart rate in bpm, mean (SD) | 71 (11) | 71 (11) | 74 (12) | 71 (11) | 70 (11) | 70 (10) |

bpm=beats per minute; CVD=cardiovascular disease; IQR=interquartile range; PAEE=physical activity energy expenditure; SD=standard deviation

**Table S7** Baseline characteristics of men not in paid employment in UK Biobank.

|  | Retired | Caring for home/family | Unable to work due to illness | Unemployed | Unpaid work | Student |
| --- | --- | --- | --- | --- | --- | --- |
| n (%) | 65,044 (83) | 1,069 (1) | 7,191 (9) | 4,609 (6) | 579 (1) | 377 (0) |
| Working hours per week, median (IQR) | 0 | 0 | 0 | 0 | 0 | 0 |
| Age in years at baseline, mean (SD) | 64 (4) | 52 (7) | 56 (7) | 53 (7) | 56 (8) | 48 (7) |
| White ethnicity, % | 98 | 88 | 94 | 87 | 88 | 70 |
| *Highest educational level* |  |  |  |  |  |  |
| No qualification, % | 24 | 19 | 44 | 23 | 9 | 6 |
| Any other qualification, % | 45 | 50 | 44 | 52 | 40 | 31 |
| Degree or above, % | 31 | 31 | 12 | 26 | 51 | 63 |
| Townsend index (higher for more deprived), median (IQR) | -2.6 (-3.9 to -0.4) | -.01 (-2.8 to 3.0) | 1.6 (-1.6 to 4.4) | 0.5 (-2.5 to 3.9) | -0.4 (-3.0 to 2.8) | 0.7 (-2.2 to 4.1) |
| *Household income before tax* |  |  |  |  |  |  |
| Prefer not to answer, % | 9 | 8 | 7 | 8 | 10 | 7 |
| Do not know, % | 2 | 7 | 10 | 9 | 6 | 8 |
| <£18,000, % | 30 | 40 | 65 | 53 | 37 | 42 |
| £18,000-£30,999, % | 30 | 18 | 11 | 14 | 17 | 19 |
| £31,000-£51,999, % | 19 | 13 | 4 | 9 | 15 | 15 |
| £52,000-£100,000, % | 8 | 10 | 2 | 5 | 10 | 7 |
| >£100,000, % | 1 | 4 | 0 | 1 | 4 | 2 |
| *Smoking status* |  |  |  |  |  |  |
| Never, % | 43 | 43 | 30 | 44 | 51 | 60 |
| Previous, % | 48 | 34 | 39 | 31 | 32 | 25 |
| Current, % | 9 | 23 | 31 | 25 | 17 | 15 |
| *Alcohol use status* |  |  |  |  |  |  |
| Never, % | 2 | 6 | 4 | 5 | 4 | 15 |
| Previous, % | 3 | 7 | 13 | 6 | 7 | 9 |
| Current, % | 94 | 87 | 83 | 90 | 89 | 76 |
| Fruit/vegetable score, median (IQR) | 1 (1 to 2) | 1 (0 to 2) | 1 (0 to 2) | 1 (0 to 2) | 1 (1 to 2) | 1 (1 to 2) |
| Red/processed meat score, median (IQR) | 1 (1 to 1) | 1 (1 to 1) | 1 (1 to 2) | 1 (1 to 1) | 1 (1 to 1) | 1 (1 to 2) |
| Adds salt to food, % | 41 | 42 | 45 | 45 | 39 | 40 |
| Consumes oily fish, % | 30 | 36 | 31 | 34 | 32 | 36 |
| Non-work PAEE (kJ/kg/day), mean (SD) | 42 (3) | 42 (4) | 40 (3) | 41 (3) | 42 (3) | 42 (4) |
| Parental history of CVD or cancer, % | 73 | 68 | 72 | 65 | 65 | 55 |
| Blood pressure or cholesterol medication, % | 49 | 26 | 53 | 28 | 29 | 16 |
| Diagnosis of diabetes or insulin prescription, % | 9 | 7 | 16 | 9 | 6 | 5 |
| Prevalent CVD or cancer at baseline, % | 25 | 12 | 35 | 12 | 14 | 5 |
| *Body mass index* |  |  |  |  |  |  |
| <25 kg/m2, % | 25 | 30 | 21 | 27 | 31 | 30 |
| 25-30 kg/m2, % | 51 | 44 | 38 | 44 | 47 | 50 |
| >30 kg/m2, % | 24 | 26 | 41 | 28 | 22 | 20 |
| Resting heart rate in bpm, mean (SD) | 68 (12) | 70 (13) | 74 (14) | 71 (13) | 69 (12) | 70 (11) |

bpm=beats per minute; CVD=cardiovascular disease; IQR=interquartile range; PAEE=physical activity energy expenditure; SD=standard deviation

**Table S8** Distribution of participants across occupational physical activity categories within strata of standard occupational code in women (n=138,062) and men (n=126,362) in UK Biobank.

|  |  | Occupational physical activity category | | | | | |  |
| --- | --- | --- | --- | --- | --- | --- | --- | --- |
| Sex | Standard occupational code strata | No heavy manual, no standing/walking | No heavy manual, some standing/walking | No heavy manual, mostly standing/walking | Some heavy manual, some standing/walking | Some heavy manual, mostly standing/walking | Mostly heavy manual, mostly standing/walking | Total |
| Women | Managers and Senior Officials | 9,410 (54%) | 4,054 (23%) | 885 (5%) | 1,038 (6%) | 1,261 (7%) | 824 (5%) | 17,472 (100%) |
|  | Professional | 10,242 (34%) | 8,850 (29%) | 6,672 (22%) | 1,391 (5%) | 2,576 (8%) | 579 (2%) | 30,310 (100%) |
|  | Associate Professional and Technical | 10,082 (36%) | 6,362 (23%) | 2,477 (9%) | 2,377 (8%) | 3,897 (14%) | 2,901 (10%) | 28,096 (100%) |
|  | Administrative and Secretarial | 19,186 (58%) | 8,444 (26%) | 1,709 (5%) | 1,981 (6%) | 1,192 (4%) | 473 (1%) | 32,985 (100%) |
|  | Skilled Trades | 153 (7%) | 140 (6%) | 155 (7%) | 175 (8%) | 661 (29%) | 958 (43%) | 2,242 (100%) |
|  | Personal Care and Service | 899 (7%) | 2,230 (17%) | 2,300 (17%) | 1,282 (10%) | 3,502 (26%) | 3,027 (23%) | 13,240 (100%) |
|  | Sales and Customer Service | 1,417 (21%) | 748 (11%) | 1,141 (17%) | 311 (5%) | 1,975 (29%) | 1,186 (17%) | 6,778 (100%) |
|  | Process, Plant and Machine Operatives | 402 (30%) | 170 (13%) | 108 (8%) | 176 (13%) | 208 (15%) | 289 (21%) | 1,353 (100%) |
|  | Elementary | 245 (4%) | 208 (4%) | 857 (15%) | 189 (3%) | 1,543 (28%) | 2,544 (46%) | 5,586 (100%) |
| Men | Managers and Senior Officials | 12,343 (44%) | 7,621 (27%) | 1,375 (5%) | 2,752 (10%) | 2,506 (9%) | 1,447 (5%) | 28,044 (100%) |
|  | Professional | 14,058 (43%) | 10,681 (33%) | 3,575 (11%) | 1,667 (5%) | 1,816 (6%) | 665 (2%) | 32,462 (100%) |
|  | Associate Professional and Technical | 7,471 (39%) | 5,036 (26%) | 1,442 (7%) | 2,049 (11%) | 2,085 (11%) | 1,147 (6%) | 19,230 (100%) |
|  | Administrative and Secretarial | 4,444 (54%) | 1,864 (23%) | 436 (5%) | 796 (10%) | 458 (6%) | 281 (3%) | 8,279 (100%) |
|  | Skilled Trades | 1,058 (6%) | 803 (5%) | 405 (2%) | 1,471 (9%) | 3,841 (23%) | 9,203 (55%) | 16,781 (100%) |
|  | Personal Care and Service | 197 (7%) | 357 (13%) | 415 (15%) | 317 (11%) | 823 (29%) | 726 (26%) | 2,835 (100%) |
|  | Sales and Customer Service | 530 (24%) | 315 (14%) | 275 (12%) | 172 (8%) | 513 (23%) | 443 (20%) | 2,248 (100%) |
|  | Process, Plant and Machine Operatives | 2,915 (29%) | 801 (8%) | 320 (3%) | 1,524 (15%) | 1,415 (14%) | 3,011 (30%) | 9,986 (100%) |
|  | Elementary | 308 (5%) | 468 (7%) | 599 (9%) | 494 (8%) | 1,619 (25%) | 3,009 (46%) | 6,497 (100%) |

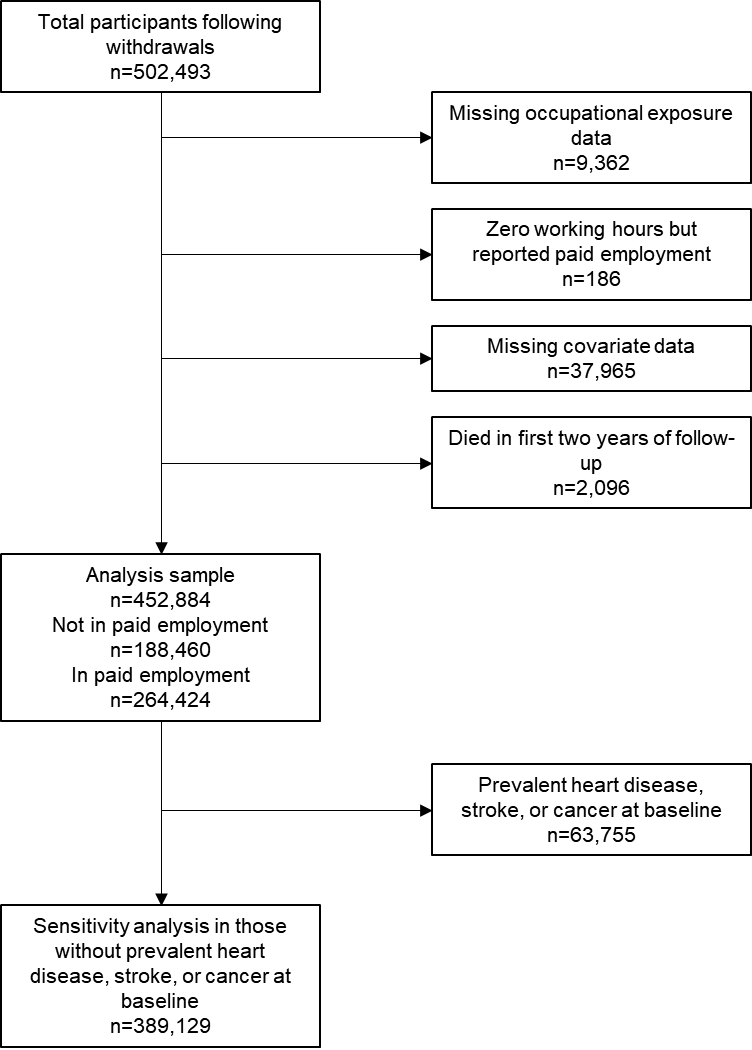

**Figure S1** Flowchart detailing participant exclusions.

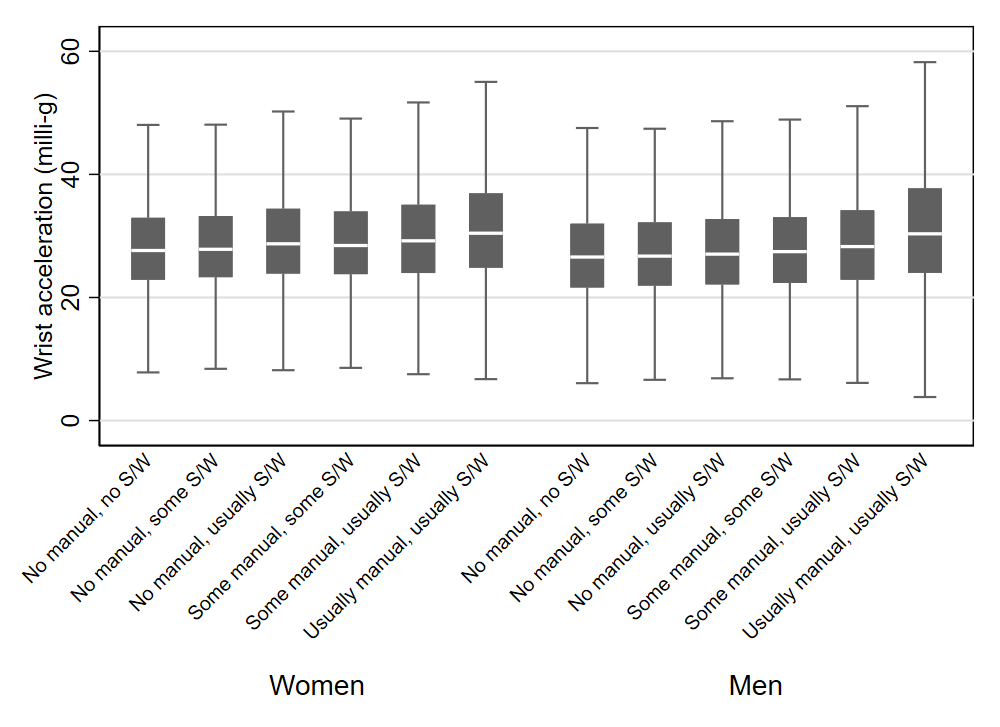

**Figure S2** Median, interquartile range, upper and lower adjacent values of average wrist acceleration in milli-g by occupational physical activity strata. Data are for those participants in the UK Biobank accelerometery sub-cohort with valid accelerometer data and in paid employment at baseline (n=63,512).

Physical activity assessment using accelerometers occurred a median 5.7 years after baseline assessment of occupational physical activity variables.

S/W=standing/walking.

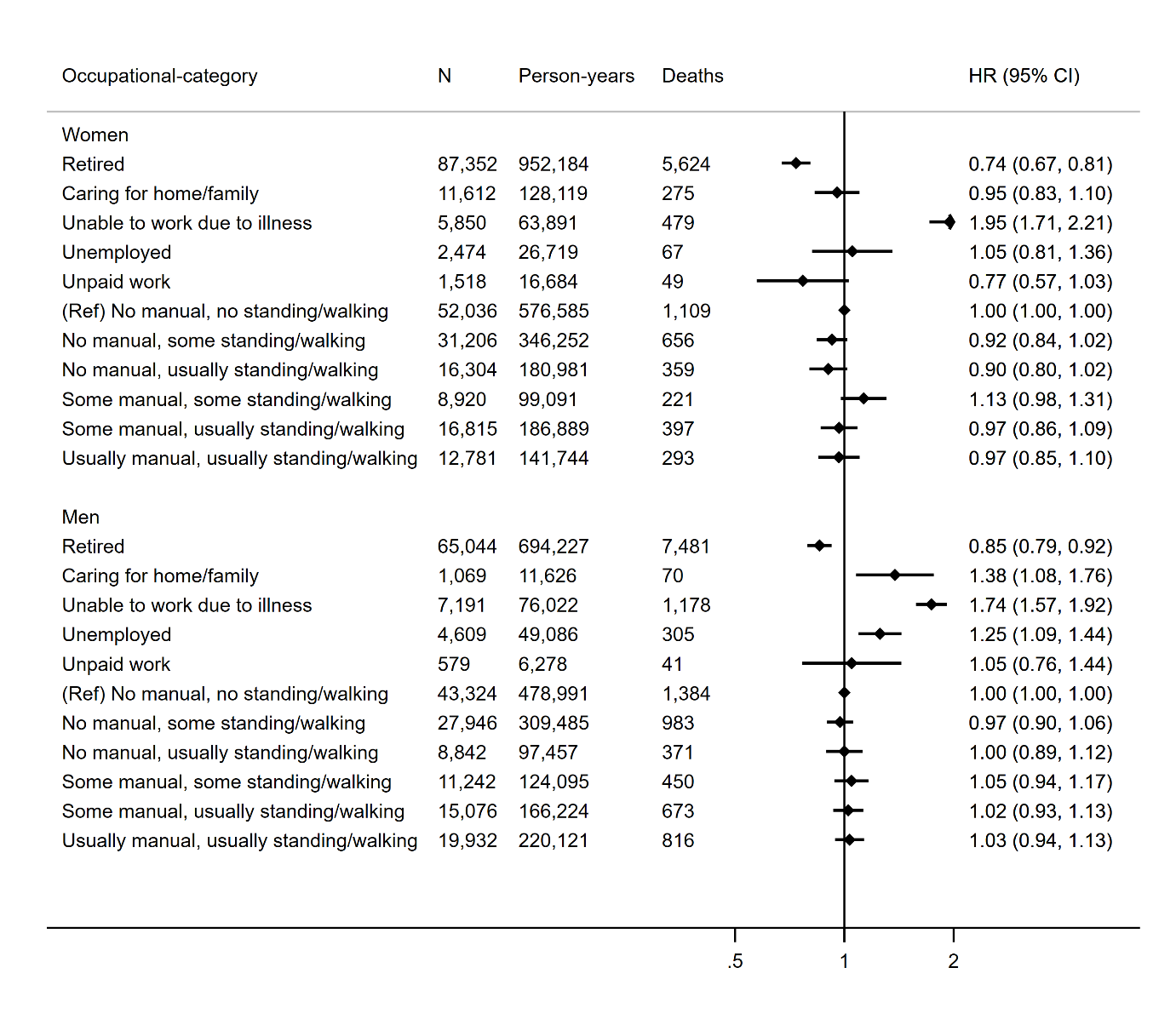

**Figure S3** Hazard ratio (HR) and 95% confidence interval (CI) of all-cause mortality by occupational category. Reference group is “no manual, no standing/walking”.

Models adjusted for age (underlying timescale), ethnicity, Townsend deprivation index, highest educational level (stratified baseline hazard), annual household income (stratified baseline hazard), alcohol consumption, smoking, salt added to food, oily fish intake, fruit and vegetable intake (stratified baseline hazard), processed and red meat intake, non-occupational physical activity energy expenditure, parental history of cancer or CVD, use of blood pressure or cholesterol lowering medications, doctor-diagnosed diabetes or treatment with insulin, baseline prevalent coronary heart disease, stroke, or cancer, body mass index, resting heart rate.

| 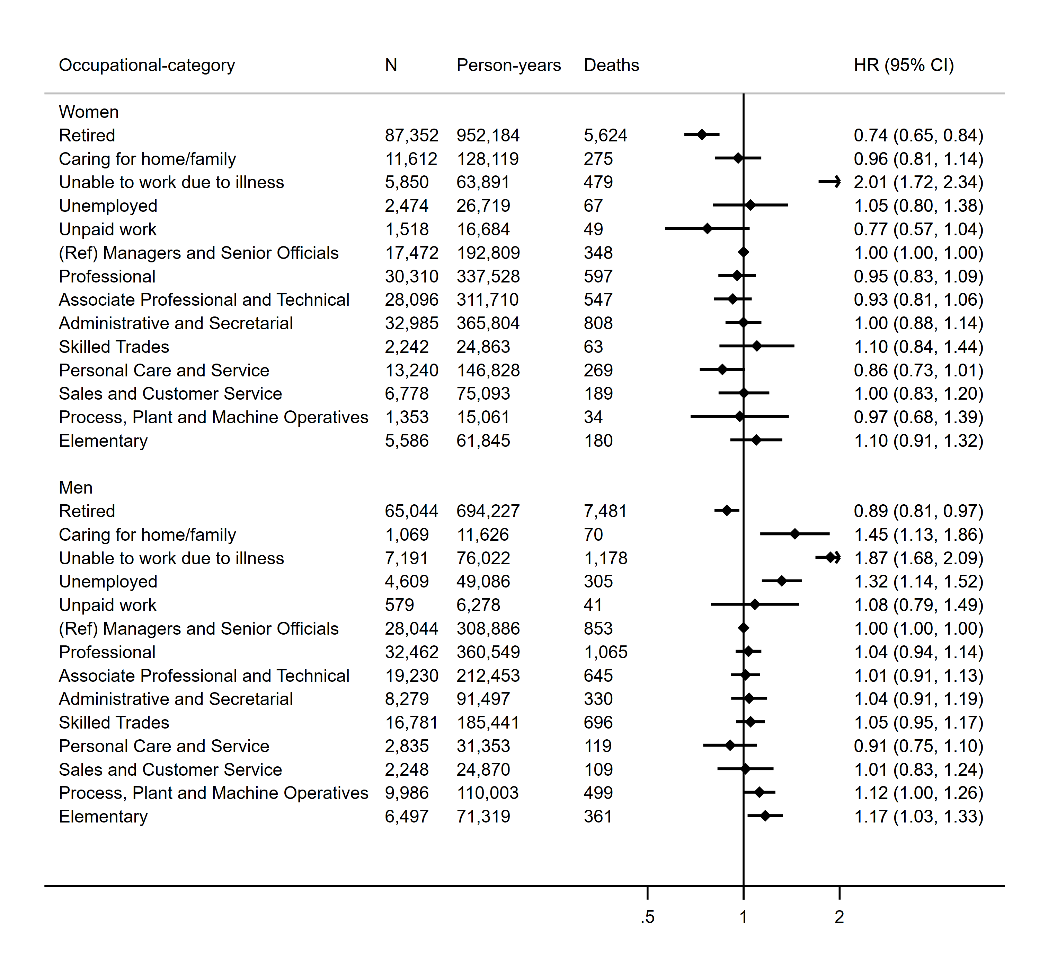 | 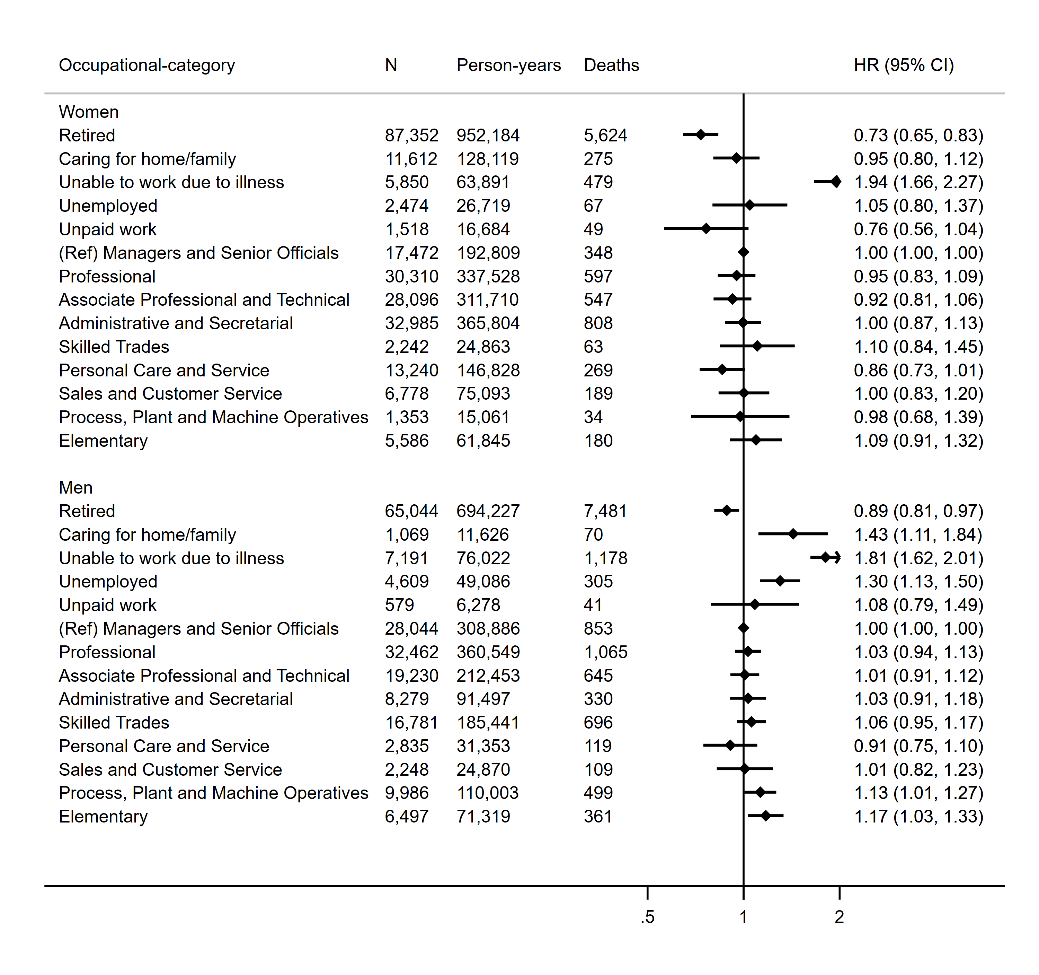 |
| --- | --- |
| **Figure S4** Hazard ratio (HR) and 95% confidence interval (CI) of all-cause mortality by occupational category for Model 1 (left) and Model 2 (right). Reference group is “Managers and Senior Officials”.  Model 1 adjusted for age (underlying timescale), ethnicity, Townsend deprivation index, highest educational level (stratified baseline hazard), annual household income (stratified baseline hazard), working hours per week, alcohol consumption, smoking, salt added to food, oily fish intake, fruit and vegetable intake (stratified baseline hazard), processed and red meat intake, non-occupational physical activity energy expenditure, parental history of cancer or CVD, use of blood pressure or cholesterol lowering medications, doctor-diagnosed diabetes or treatment with insulin, baseline prevalent coronary heart disease, stroke or cancer.  Model 2 additionally adjusted for body mass index, resting heart rate. | |

| 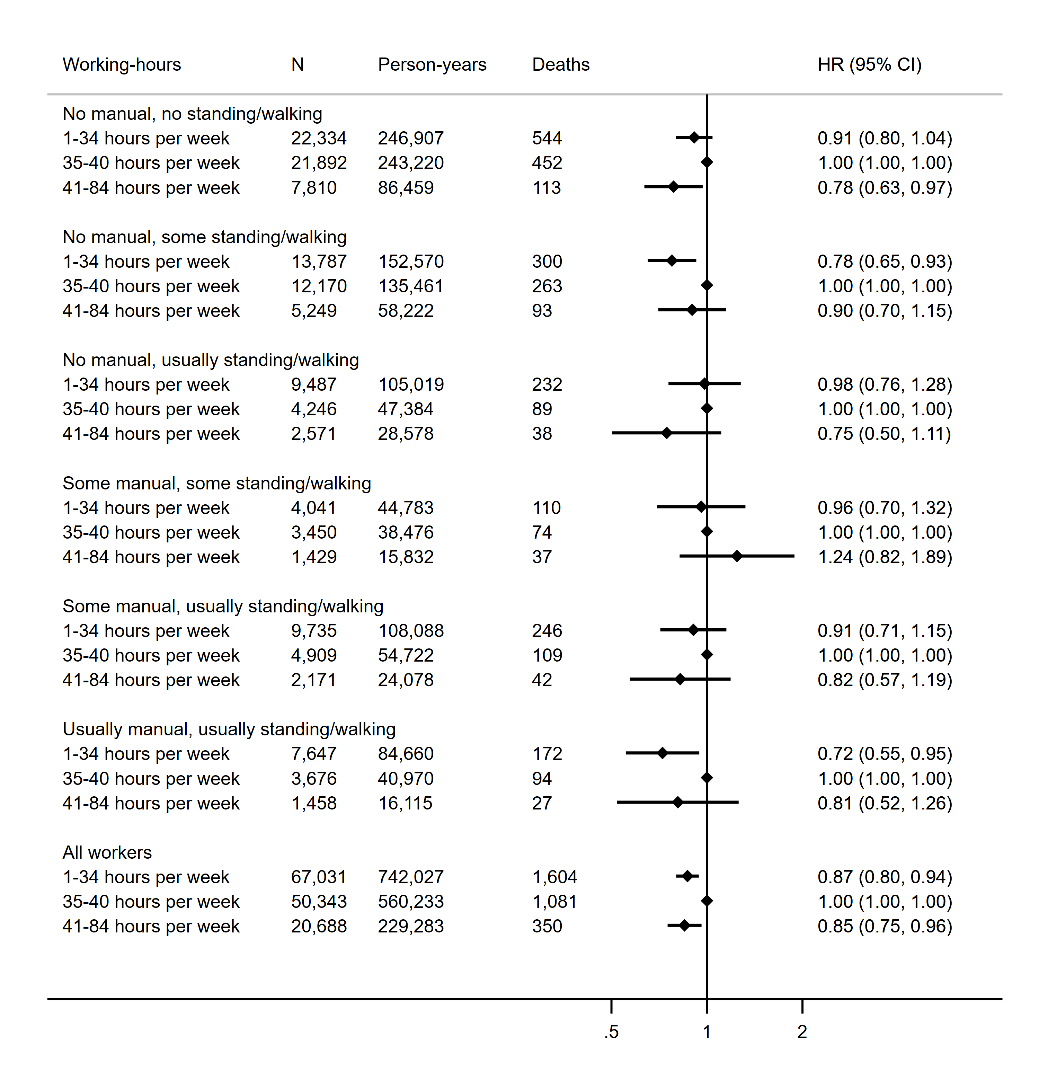 | 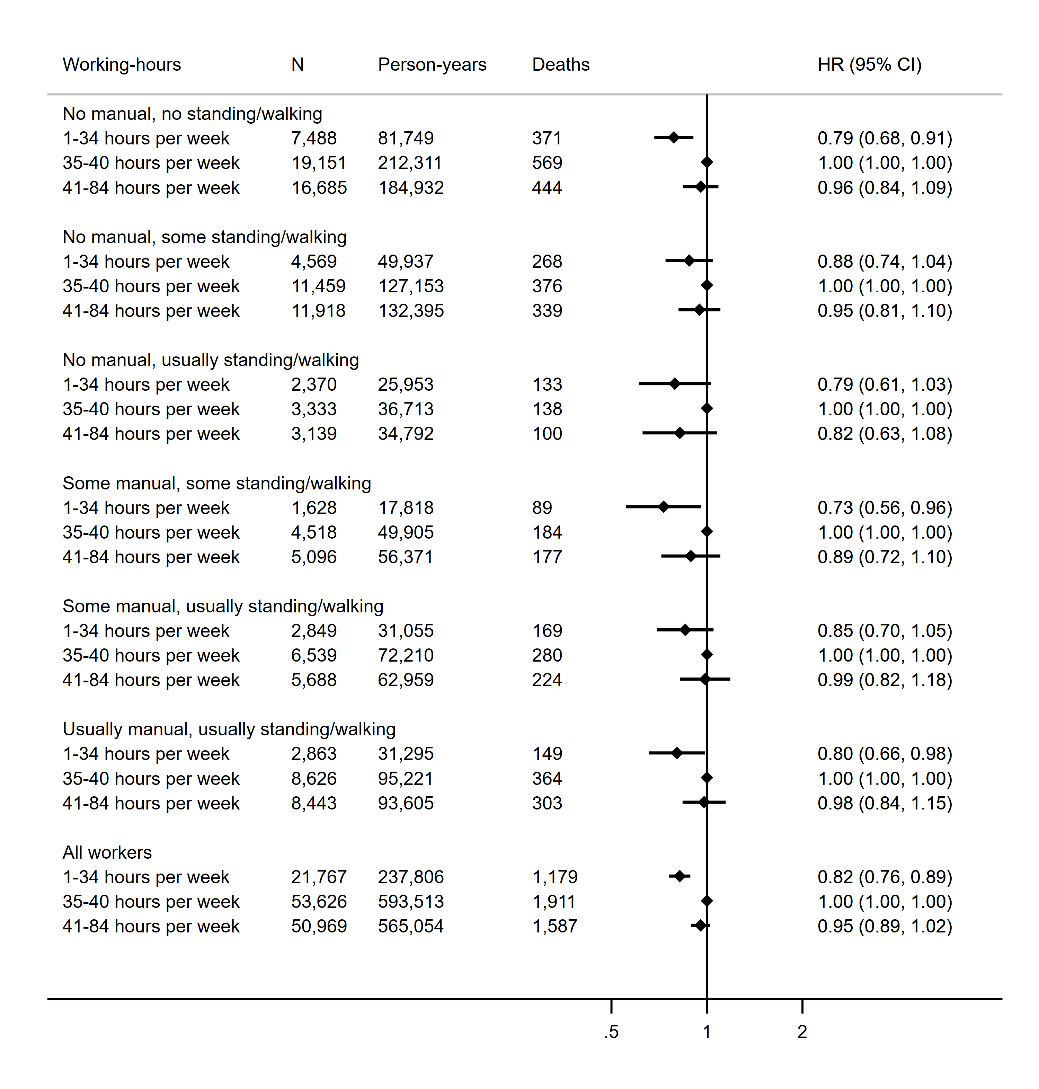 |
| --- | --- |
| **Figure S5** Hazard ratio (HR) and 95% confidence interval (CI) of all-cause mortality by tertile of working hours per week across occupational physical activity strata in women (left) and men (right). Reference group is “35-40 hours per week”.  Models are adjusted for age (underlying timescale), ethnicity, Townsend deprivation index, highest educational level (stratified baseline hazard), annual household income (stratified baseline hazard), alcohol consumption, smoking, salt added to food, oily fish intake, fruit and vegetable intake (stratified baseline hazard), processed and red meat intake, non-occupational physical activity energy expenditure, parental history of cancer or CVD, use of blood pressure or cholesterol lowering medications, doctor-diagnosed diabetes or treatment with insulin, baseline prevalent coronary heart disease, stroke, or cancer, body mass index, resting heart rate. | |
| 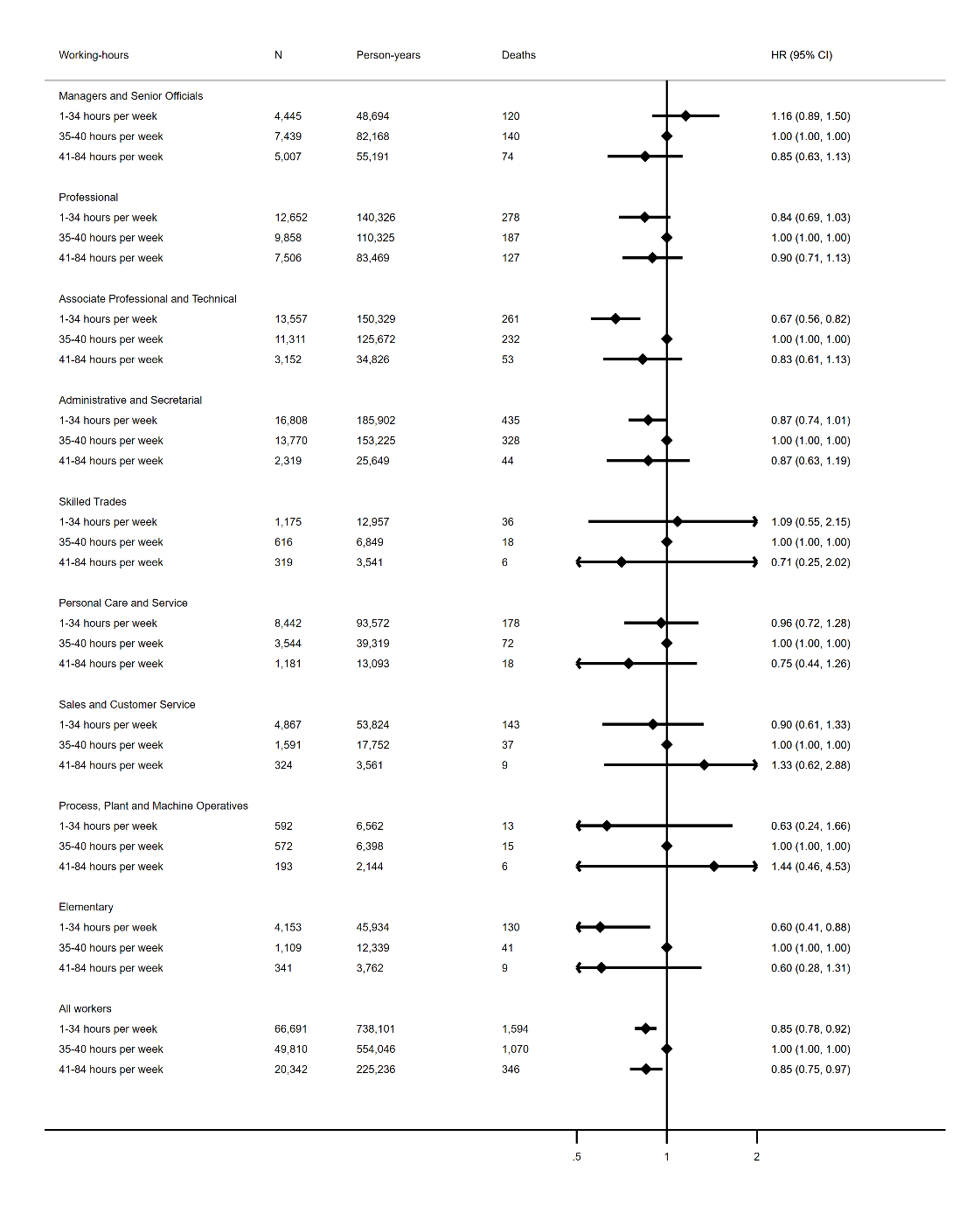 | 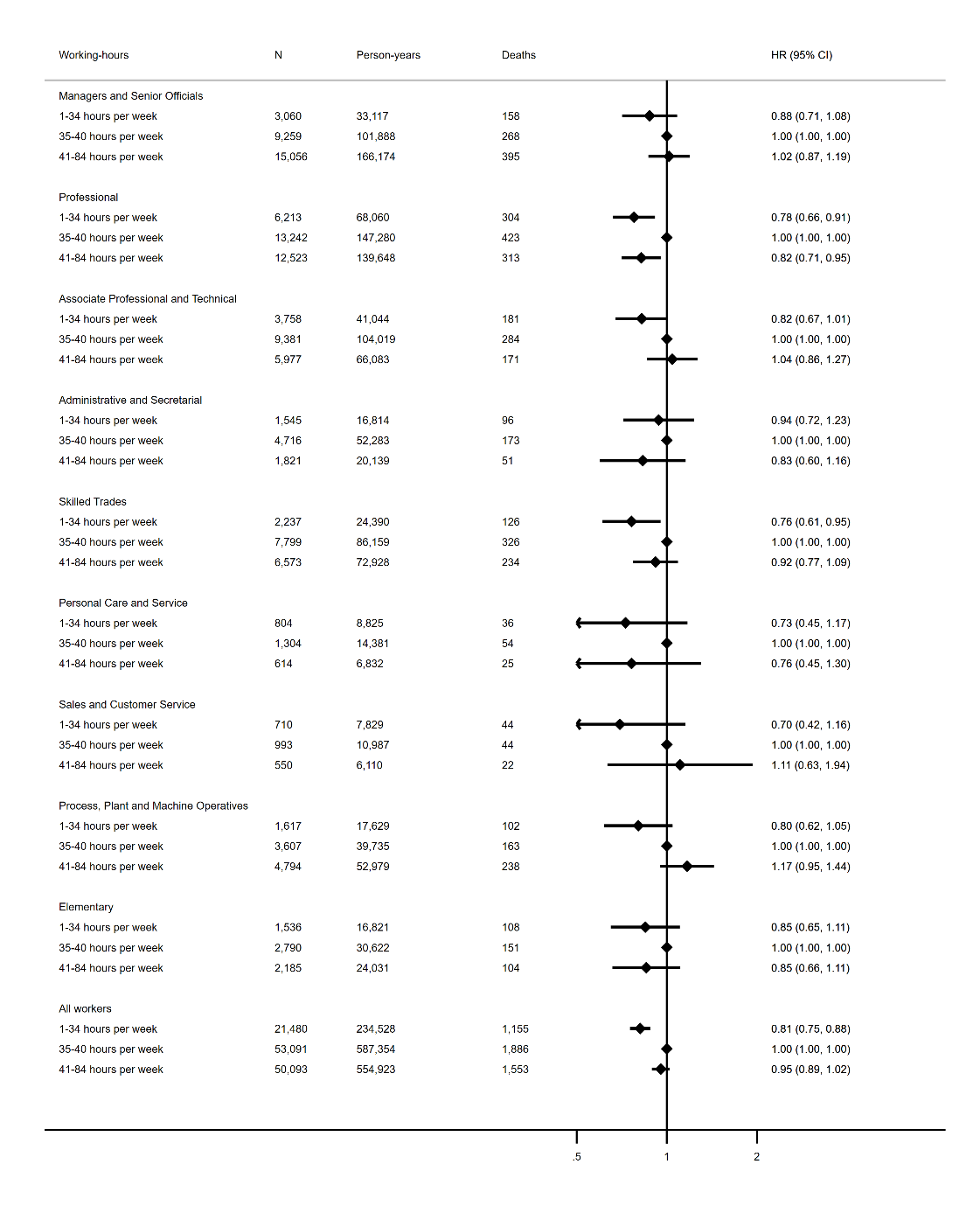 |
| **Figure S6** Hazard ratio (HR) and 95% confidence interval (CI) of all-cause mortality by tertile of working hours per week across standard occupational code strata for women (left) and men (right). Reference group is “35-40 hours per week”.  Models are adjusted for age (underlying timescale), ethnicity, Townsend deprivation index, highest educational level (stratified baseline hazard), annual household income (stratified baseline hazard), alcohol consumption, smoking, salt added to food, oily fish intake, fruit and vegetable intake (stratified baseline hazard), processed and red meat intake, non-occupational physical activity energy expenditure, parental history of cancer or CVD, use of blood pressure or cholesterol lowering medications, doctor-diagnosed diabetes or treatment with insulin, baseline prevalent coronary heart disease, stroke or cancer, body mass index, resting heart rate. | |

| 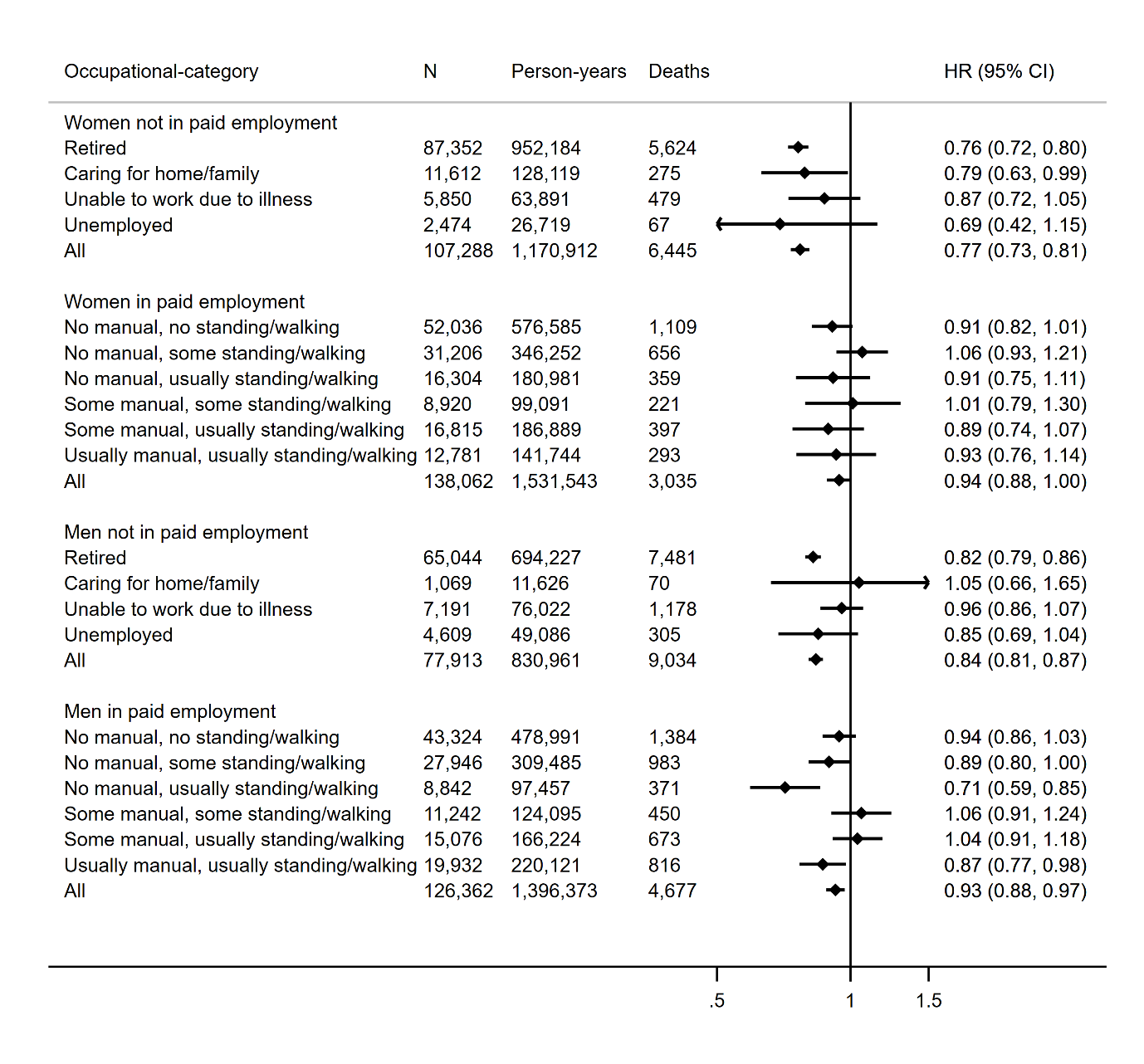 |
| --- |
| **Figure S7** Hazard ratio (HR) and 95% confidence interval (CI) of all-cause mortality per 5 kJ/day/kg of non-occupational physical activity energy expenditure across occupational strata.  Models adjusted for age (underlying timescale), ethnicity, Townsend deprivation index, highest educational level (stratified baseline hazard), annual household income (stratified baseline hazard), working hours per week, alcohol consumption, smoking, salt added to food, oily fish intake, fruit and vegetable intake (stratified baseline hazard), processed and red meat intake, parental history of cancer or CVD, use of blood pressure or cholesterol lowering medications, doctor-diagnosed diabetes or treatment with insulin, baseline prevalent coronary heart disease, stroke, or cancer, body mass index, resting heart rate. |

| 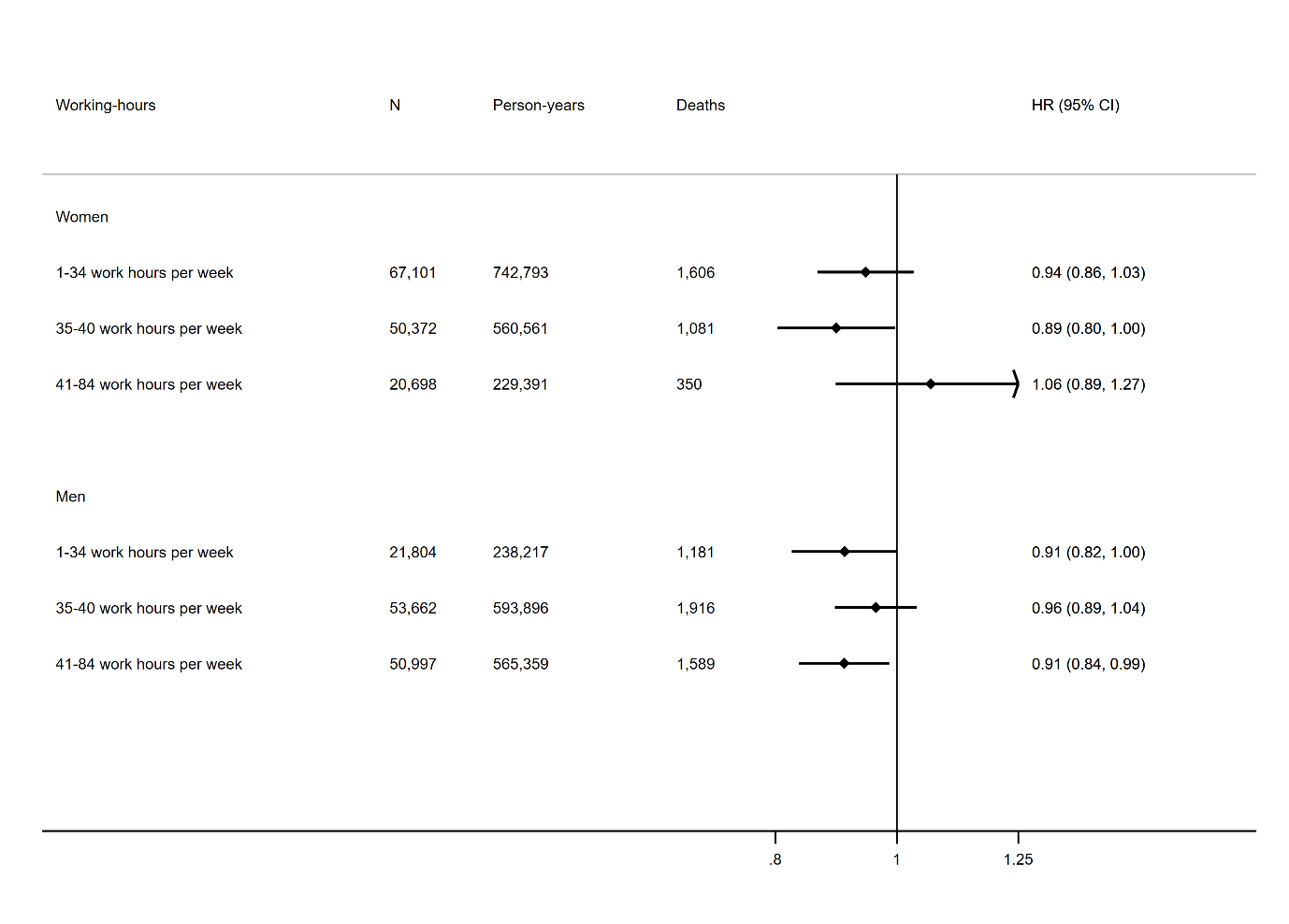 |
| --- |
| **Figure S8** Hazard ratio (HR) and 95% confidence interval (CI) of all-cause mortality per 5 kJ/day/kg of non-occupational physical activity energy expenditure across tertiles of working hours per week.  Models adjusted for age (underlying timescale), ethnicity, Townsend deprivation index, highest educational level (stratified baseline hazard), annual household income (stratified baseline hazard), alcohol consumption, smoking, salt added to food, oily fish intake, fruit and vegetable intake (stratified baseline hazard), processed and red meat intake, parental history of cancer or CVD, use of blood pressure or cholesterol lowering medications, doctor-diagnosed diabetes or treatment with insulin, baseline prevalent coronary heart disease, stroke or cancer, occupational physical activity category, body mass index, resting heart rate. |
